# Identifying Metabolomic and Proteomic Biomarkers for Age-Related Morbidity in a Population-Based Cohort - the Cooperative Health Research in South Tyrol (CHRIS) study

**DOI:** 10.1101/2024.07.15.24310410

**Authors:** Essi Hantikainen, Christian X. Weichenberger, Nikola Dordevic, Vinicius Verri Hernandes, Luisa Foco, Martin Gögele, Roberto Melotti, Cristian Pattaro, Markus Ralser, Fatma Amari, Vadim Farztdinov, Michael Mülleder, Peter P. Pramstaller, Johannes Rainer, Francisco S. Domingues

**Affiliations:** Institute for Biomedicine, Eurac Research, Bolzano, Italy; Department of Food Chemistry and Toxicology, University of Vienna, Vienna, Austria; Department of Biochemistry, Charité – Universitätsmedizin Berlin, Corporate Member of Freie Universität Berlin and Humboldt-Universität zu Berlin, Berlin, Germany; Core Facility, High-Throughput Mass Spectrometry, Charité, Universitätsmedizin Berlin, Corporate Member of Freie Universität Berlin and Humboldt-Universität Zu Berlin, Berlin, Germany

**Keywords:** Health Status, Metabolomics, Proteomics, Comorbidity, Aging, CHRIS study

## Abstract

Identifying biomarkers able to discriminate individuals on different health trajectories is crucial to understand the molecular basis of age-related morbidity. We investigated multi-omics signatures of general health and organ-specific morbidity, as well as their interconnectivity. We examined cross-sectional metabolome and proteome data from 3,142 adults of the Cooperative Health Research in South Tyrol (CHRIS) study, an Alpine population study designed to investigate how human biology, environment, and lifestyle factors contribute to people’s health over time. We had 174 metabolites and 148 proteins quantified from fasting serum and plasma samples. We used the Cumulative Illness Rating Scale (CIRS) Comorbidity Index (CMI), which considers morbidity in 14 organ systems, to assess health status (any morbidity vs. healthy). Omics-signatures for health status were identified using random forest (RF) classifiers. Linear regression models were fitted to assess directionality of omics markers and health status associations, as well as to identify omics markers related to organ-specific morbidity.

Next to age, we identified 21 metabolites and 10 proteins as relevant predictors of health status and results confirmed associations for serotonin and glutamate to be age-independent. Considering organ-specific morbidity, several metabolites and proteins were jointly related to endocrine, cardiovascular, and renal morbidity. To conclude, circulating serotonin was identified as a potential novel predictor for overall morbidity.

## 1. Introduction

Non-communicable diseases (NCDs) are the leading cause of morbidity and premature mortality globally^1^. Age itself is the leading predictor for most NCDs: NCD prevalence increases with age and multiple diseases tend to cluster among older individuals^2^. Common biological processes triggered by molecular damage and modified by cellular and systemic responses drive biological aging and modify risks for multiple diseases in a tissue-, organ- and system-specific manner^3^. In turn, these health outcomes feed back into the underlying biological processes impacting the rate of aging and enhancing the risk for further disease^4^. It is therefore important to enhance our understanding of the molecular basis of age-related diseases to improve measures for disease prevention and general health^5^. Omics-based biomarkers provide insights into the molecular processes driving functional decline, they also help monitoring health trajectories, age-related physiological decline and disease onset^6^. Such biomarkers can also support the development of prevention strategies targeting those processes and provide surrogate endpoints in intervention studies^5^. The goal is early identification of individuals at higher risk of diseases, who will benefit most from such preventive interventions^7^. Protein biomarkers have the advantage of being direct biological effectors of the underlying genomic background^7^. Serum metabolomics on the other hand provides a snapshot of general physiological state of an organism, and is influenced by genetic, epigenetic, and environmental factors^8,9^. For instance, Tanaka et al. (2020)^7^ identified a proteomic signature of aging involving 76 proteins and predicting accumulation of chronic diseases and all-cause mortality. You et al. (2023)^10^ developed a disease specific proteomic risk score, which stratified the risk for 45 common disease conditions, resulting into an equivalent predictive performance over established clinical indicators for almost all endpoints. Similarly, Gadd et al. (2024)^11^ demonstrated the utility of proteomic scores in predicting several 10-year incident outcomes beyond factors, such as age, sex, lifestyle and clinically relevant biomarkers, showing the relevance of early proteomic contributions to major age-related diseases. Pietzner et al. (2021)^12^ used untargeted metabolomics to investigate signatures of multimorbidity and found that 420 metabolites are shared between at least two chronic diseases. A recent work demonstrated the potential of metabolomic profiles as a multi-disease assay to inform on the risk of many common diseases simultaneously. For 10-year outcome prediction of 15 selected endpoints a combination of age, sex and the metabolomic state was equal or outperformed established predictors^13^.

Advances in different omics technologies and computing capabilities have also enabled the integration of multi-omics data to capture the complex molecular interplay of health and disease^14^. In TwinsUK, using data from 510 women, Zierer et al. (2016)^15^ integrated four high-throughput omics datasets and demonstrated the interconnectivity of age-related diseases by highlighting molecular markers of the aging process, which might drive disease comorbidities.

Here, we aimed to identify multi-omics signatures of general health among adult individuals using cross sectional data from the population-based Cooperative Health Research in South Tyrol (CHRIS) study, an Alpine population study designed to investigate how human biology, environment, and lifestyle factors contribute to people’s health over time. We specifically included targeted serum metabolomics^16^ and plasma proteomics data^17^. The Cumulative Illness Rating Scale (CIRS) based Comorbidity Index (CMI)^18^, reflecting disease status and severity in 14 relevant organ systems, was used to assess health status by classifying individuals into having any morbidity (CMI≥1) or being healthy (CMI=0). We then applied predictive models using a random forest classifier to determine health status. The analyses were complemented by multiple linear regression models investigating associations and inter-dependencies of individual proteins or metabolites with morbidity assessed by each organ-specific CIRS domains, such as the endocrine-metabolic or the renal domains.

## 2. Results

The main analytic sample consisted of n=3,142 adult individuals from the CHRIS study with available metabolomics and proteomics data. The AbsoluteIDQ® p180 kit from Biocrates (Biocrates Life Sciences AG, Innsbruck, Austria) was used for metabolite quantification in fasting serum samples^16^. The high abundance plasma proteome was determined using the Scanning SWATH mass spectrometry-based approach^17^. We first present the study sample’s main characteristics by health status (any morbidity vs. healthy) and the relationships between the co-occurring comorbidities. Next, we provide findings from the random forest (RF) analysis. The analyses were complemented by the integration of multiple linear regression models to better characterize actual associations of each significant feature from the RF analysis with health status. Finally, we investigated associations of omics signatures with organ specific morbidity using linear regression models.

### 2.1 Health status and characteristics of the study sample

The characteristics for the main analytic sample are presented in **Table 1**. The CIRS organ domains most completely described by the available data (completeness≥50%) were the hypertension, cardiac, respiratory, neurological, renal, vascular, endocrine-metabolic, hepatic and psychiatric/behavioral domains (**Table 1**). The remaining domains had <50% completeness and in general a lower proportion of unhealthy individuals was observed for these domains (**Table 1**). Among all individuals, 56% (n=1,751) were affected by at least one morbidity condition (CMI≥1). As expected, these were on average older than healthy individuals (CMI=0; **Table 1**, **Figure 1)**. The top five organ domains affected by health problems were the hepatic, vascular, hypertension, endocrine-metabolic and the respiratory domain, with 27.4%, 14.9%, 12.8%, 10.3%, and 8.8% of morbidity prevalence estimates, respectively. We additionally provide the characteristics for the CHRIS cohort, regardless of available omics data, to confirm the robustness of main characteristics and estimated disease prevalences in our analytic sample. These are reported in **Supplementary Table S1 and Figure S1**.

**Figure 1.**
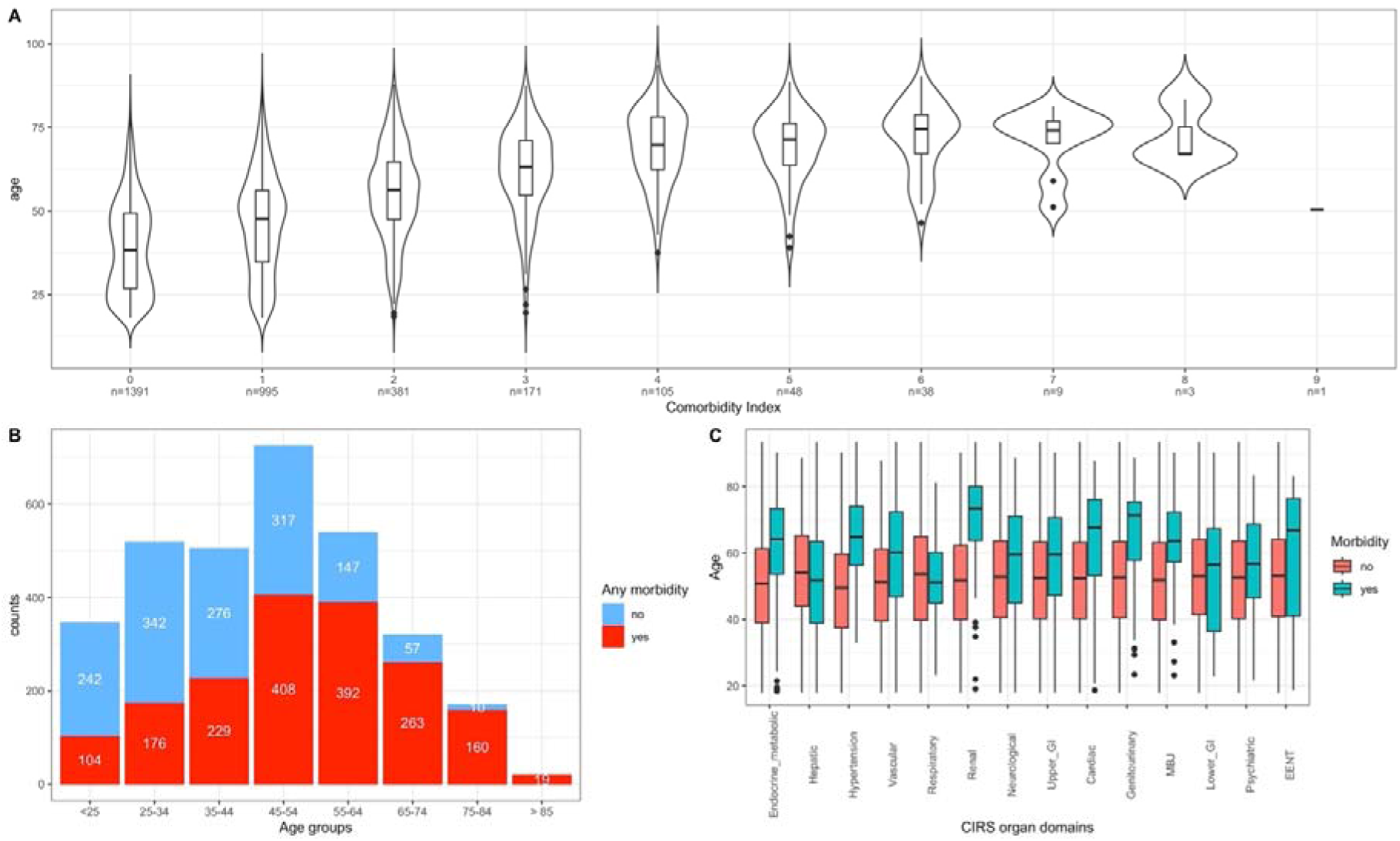
Distribution of health status and age in the CIRSHS sample. **A)** Age distribution by Comorbidity Index, which is expressed as the total number of CIRS organ domains scoring ≥2. **B)** Absolute distribution of “Any morbidity” (yes,no) by age-group. The relative proportion of red-bars within each stacked column-bar represents the age-group specific prevalence of any morbidity. **C)** Age distribution by morbidity conditions in the specific CIRS domains.

**Table 1.** Characteristics of the main analytic sample with information on health status (any morbidity vs. healthy)*^a^* and CIRS domain specific morbidity^b^.

|  | Overall | Healthy | Any morbidity | Completeness<sup>c</sup> |
| --- | --- | --- | --- | --- |
| Sample size, n (%) | **3,142** | **1,391 (44%)** | **1,751 (56%)** |  |
| Age, mean (SD) | 46.5 (16.7) | 39.0 (13.9) | 52.5 (16.3) |  |
| Comorbidity Index, mean (SD) | 1.04 (1.34) | 0 | 1.86 (1.30) |  |
| Sex, n (%) |  |  |  |  |
| Females | 1748 (55.6%) | 868 (62.4%) | 880 (50.3%) |  |
| Males | 1394 (44.4%) | 523 (37.6%) | 871 (49.7%) |  |
| Morbidity in CIRS domains, yes (%) |  |  |  |  |
| Hepatic | 861 (27.4%) |  | 861 (49.2%) | 59 |
| Vascular | 467 (14.9%) |  | 467 (26.7%) | 62 |
| Hypertension | 403 (12.8%) |  | 403 (23.0%) | 100 |
| Endocrine-Metabolic | 325 (10.3%) |  | 325 (18.6%) | 74 |
| Respiratory | 278 (8.8%) |  | 278 (15.9%) | 71 |
| Upper Gastrointestinal | 168 (5.3%) |  | 168 (9.6%) | 44 |
| Psychiatric and behavioral | 151 (4.8%) |  | 151 (8.6%) | 53 |
| MBJ<sup>d</sup> | 141 (4.5%) |  | 141 (8.1%) | 48 |
| Renal | 132 (4.2%) |  | 132 (7.5%) | 59 |
| Cardiac | 110 (3.5%) |  | 110 (6.3%) | 71 |
| Neurological | 82 (2.6%) |  | 82 (4.7%) | 66 |
| Lower Gastrointestinal | 67 (2.1%) |  | 67 (3.8%) | 44 |
| Genitourinary | 58 (1.8%) |  | 58 (3.3%) | 33 |
| EENT<sup>e</sup> | 11 (0.4%) |  | 8 (0.6%) | 32 |

<sup>a</sup>Health status was assessed through the Cumulative Illness Rating Scale (CIRS) Comorbidity Index (CMI) by classifying individuals as having any morbidity (CMI ≥ 1) or being healthy (CMI=0). <sup>b</sup>CIRS domain specific morbidity is defined as having a score ≥ 2 in the given domain. <sup>c</sup>Completeness presents the coverage of the necessary information available in the CHRIS Study with regard to the CIRS guidelines expressed in percentages. <sup>d</sup>MBJ=Musculoskeletal, bones and joints. <sup>e</sup>EENT= Ears, eyes, nose and throat.

Next, we explored relationships between the 14 CIRS domains through ordinary correspondence analysis (OCA; **Figure 2)**. We observed proximity between the hypertension, renal and endocrine domains, the cardiac and the vascular domains, and between the neurological and the psychiatric domains. To compare the robustness of the comorbidity relationships we present the OCA analysis results for the CHRIS cohort in **Supplementary Figure S2**.

**Figure 2.**
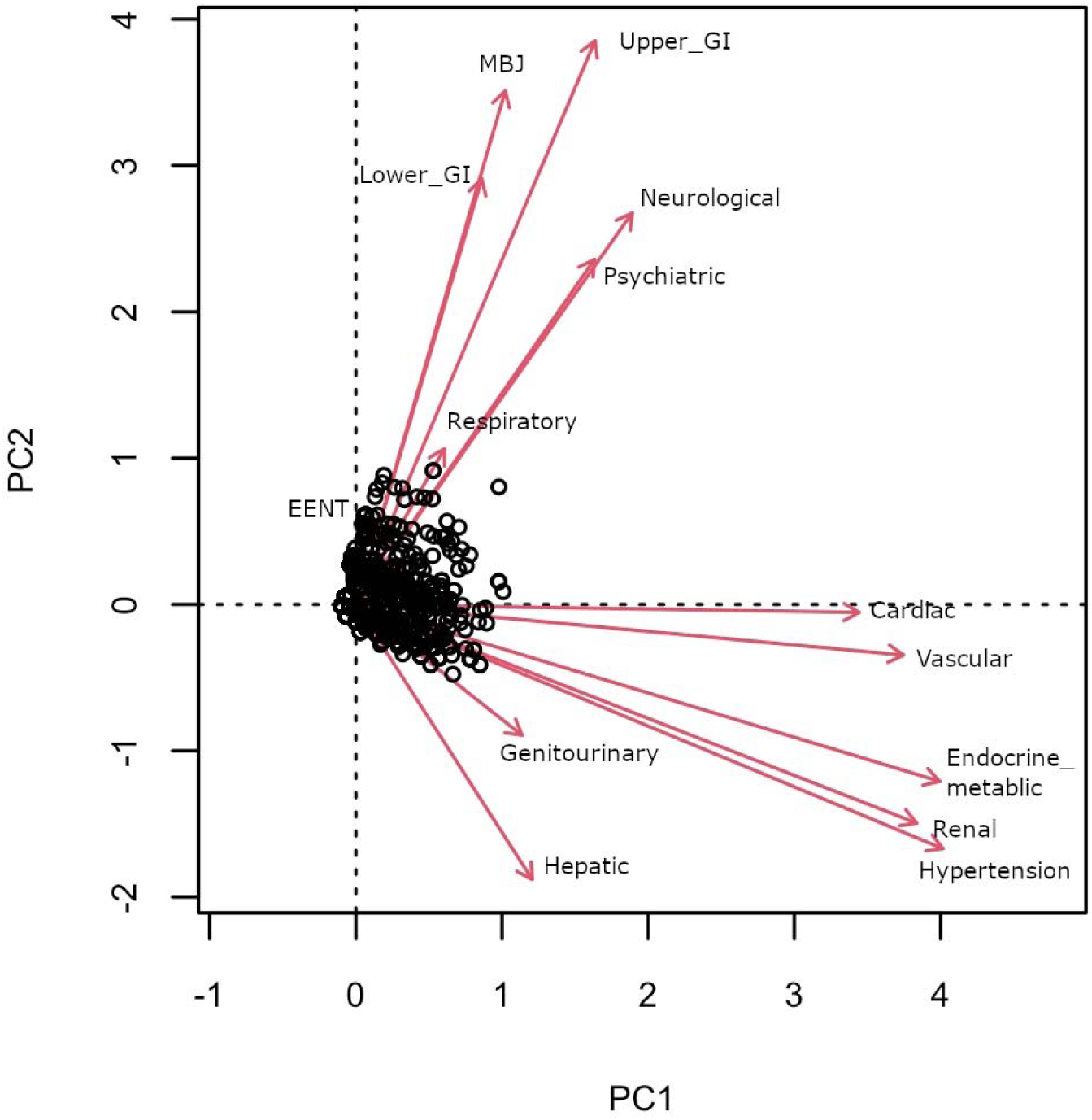
Biplot of ordinary correspondence analysis considering all subjects with available metabolomics and proteomics data, presenting relations between the 14 CIRS domains. CIRS domains with a stronger relation have longer (size consistency) and closer (direction consistency) loadings.

### 2.2 Multi omics signatures of health status

To avoid confounding of results due to the impact of medication^17^, we performed the analysis on metabolites and protein abundances adjusted for use of medications that were not considered in the CIRS definition (**Supplementary Table S2**).

For the RF analysis we built a model including age, sex, 174 metabolites and 148 proteins as predictors. Overall, 100 RF models were generated, each containing 500 trees per model, using repeated random subsampling with 80% training and 20% validation set sizes, respectively. We compared the performance measures using the area under the receiver operating curve (ROC AUC;, as well as the Matthew’s correlation coefficient (MCC) and MCC-F1. The RF model showed moderate performance (AUC=0.743, 95%Conficence Interval (CI)=0.740, 0.747; **Figure 3**; MCC and MCC-F1 are presented in **Supplementary Figure S3).** In addition, we built models that included varying sets of the predictors, which were a) age and sex, b) age, sex and metabolites, c) age, sex and proteins, and d) age, sex, metabolites, proteins. The performance comparison of these to the full model are presented in **Supplementary Text S1**. The metabolomics/proteomics based models (b,c,d) show greater performance but the differences are not statistically significant.

**Figure 3.**
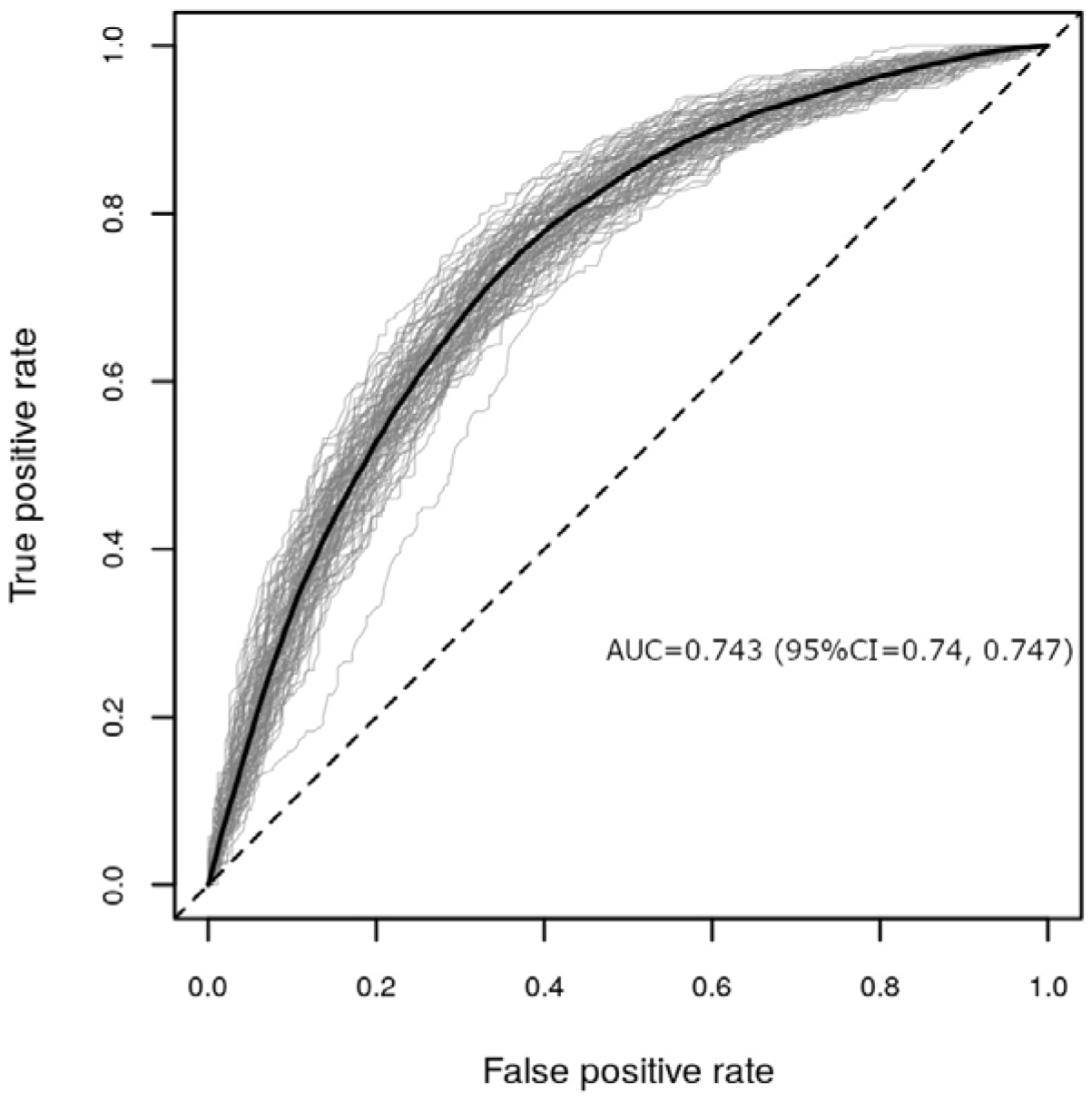
Performance evaluation of 100 random forest models including as predictors age, sex, 174 metabolites and 148 proteins to classify health status (any morbidity vs. healthy) represented as ROC AUCs. In the plots each validation run is shown as a gray line with the average curve shown in black.

We next estimated the significance of the selected individual features using permutation testing. We obtained 33 significant features, including 21 metabolites and 10 proteins, with age being the most important variable and sex the least relevant (**Figure 4A**). Among the top ten omics features were the metabolites serotonin, glutamate, hexose, three acylcarnitines (C18:1, C16:1, C16), ornithine, and the proteins CFH, A2M and IGFALS. The medication-adjusted abundance distributions of these metabolites and proteins stratified by health status are presented in **Figure 4B**. Individuals with any morbidity had lower mean abundance of serotonin, taurine and lysoPC C18:2, and higher mean abundance of all other metabolites. Regarding proteins, individuals with any morbidity had lower mean abundances of A2M, IGFALS, IGHM, and F2, and higher abundance of CFH, C4BPA, A1BG, APOH, AFM, and RBP4.

**Figure 4.**
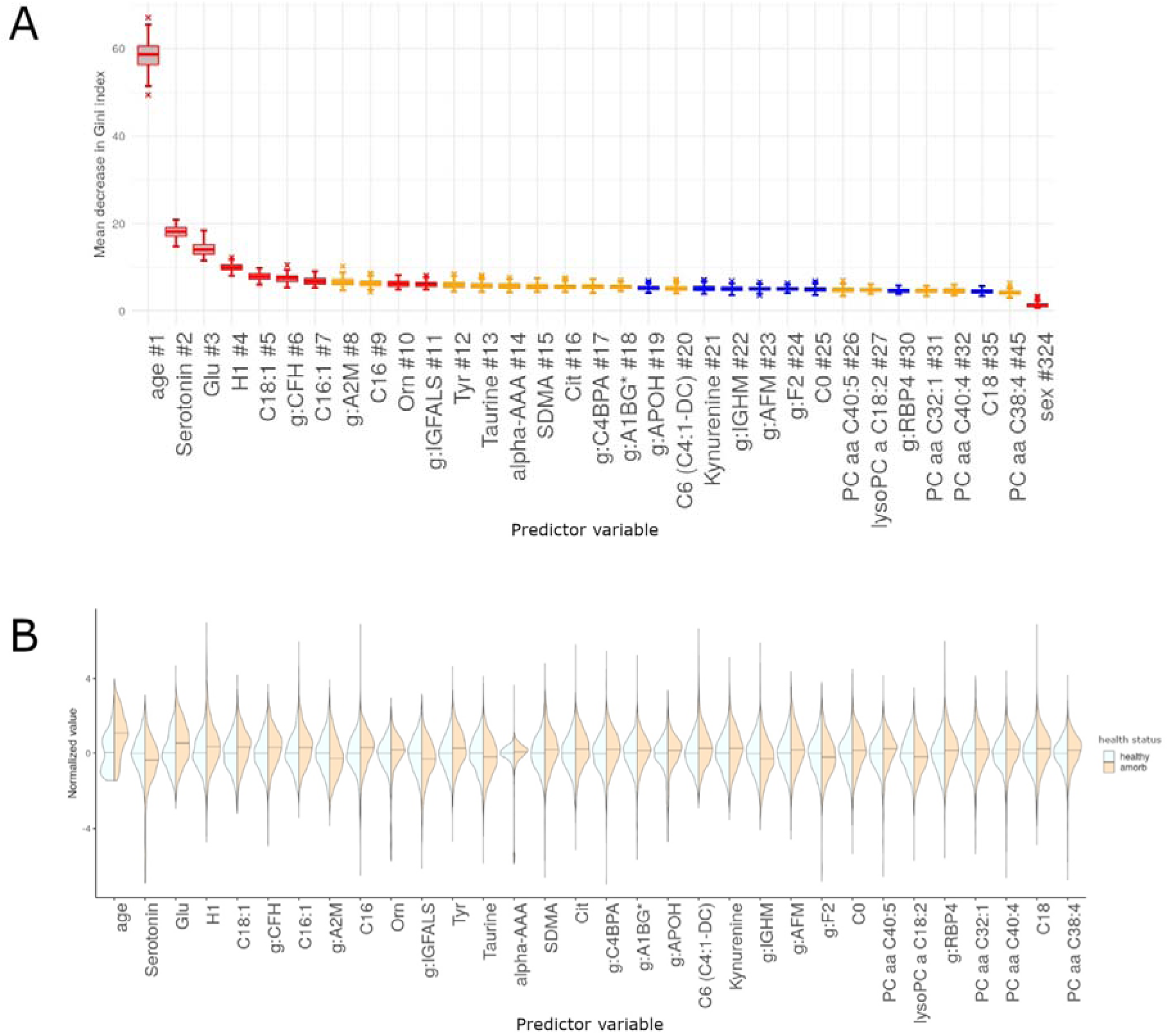
Evaluation of importance for significant features identified through the random forest models including as predictors age, sex, 174 metabolites and 148 proteins, for health status (any morbidity vs healthy). #: Rank of the specific feature. Predictor significance was estimated by permutation testing, which generates 100 background distributions, yielding 100 *p*-values for each predictor variable. Box plots are color-coded by the number of times a *p*-value was significant for such a variable (*p* <0.05): red, all 100 runs returned a significant *p*-value; orange, between 80 and 99 runs returned a significant *p*-value; blue, between 50 and 79 runs returned a significant *p*-value. Panels are restricted to features that are significant in at least 50% of all permutation runs. Density plots are scaled to have the same width, a median of zero (0) (horizontal bar) and standard deviation of one (1) for the healthy group. For comparability, the any morbidity group data have been scaled to the standard deviation of the healthy group. **A)** Mean decrease in Gini Index for significant features. **B)** Violin plots presenting scaled abundance distributions stratified by health status for significant features identified through RF, ordered by importance from left to right.

To better characterize the actual association of each significant feature from the RF analysis with health status, we performed separate regression analyses with the abundances of each identified molecular feature as the response variable and health status (any morbidity vs. healthy), age and sex as the explanatory variables, allowing us to evaluate the influence of the feature on health status independently of age or sex. Coefficients for health status were extracted from these models and are presented in **Figure 5** and **Supplementary Table S3**. Individuals with any morbidity had, on average, 22% lower mean abundance of serotonin and 12% higher abundance of serum glutamate. Twelve other features (C18:1, C16:1, lyso PC a C18:2, PC aa C32:1, tyrosine, taurine, hexose, kynurenine, AFM, CFH, RBP4, and A1BG) passed the multiple-testing correction, however, the observed differences in abundances did fall within the range of the technical variability and were thus not considered significant. Given that age was the strongest predictor for health status in the RF model, we further investigated and compared the coefficients for age and health association from the regression models. For some features, such as citrulline, abundances were almost entirely explained by age and the coefficient for the association with health status from the regression model was only very small, and not significant. For others, such as serotonin and glutamate, associations with health status were strong, even in these age-adjusted models, suggesting an age-independent association of these features with health.

**Figure 5.**
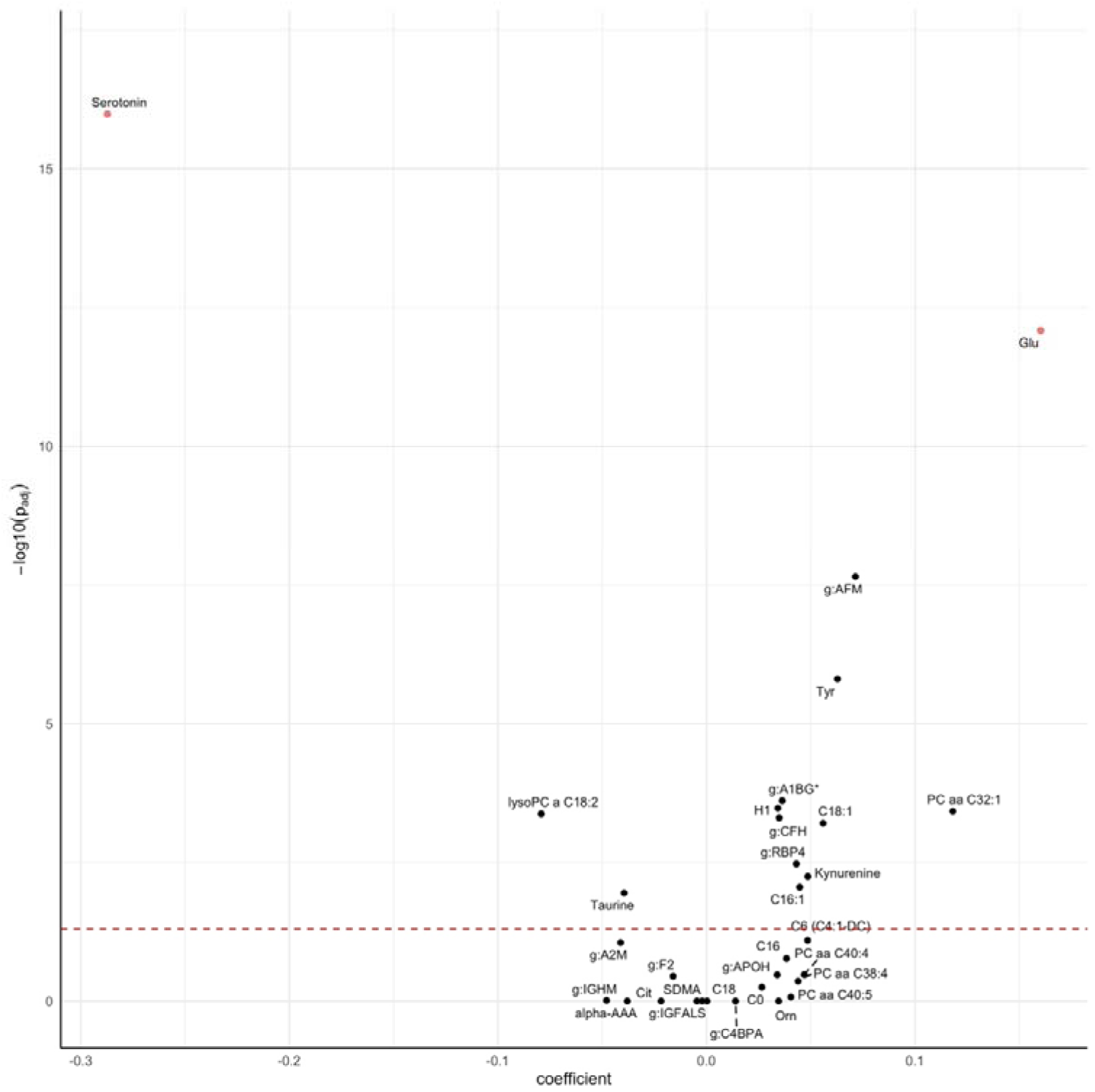
Volcano plot for the differential abundance of metabolomic and proteomic features of health status (any morbidity vs. healthy). Coefficients represent the log2□difference in average concentrations between the groups.

### 2.3 Metabolomic and proteomic signatures related to CIRS domain specific morbidity

We further evaluated morbidity markers, implementing separate regression models for medication adjusted abundances of all 174 metabolites and 148 proteins with each CIRS domain as well as age and sex as explanatory variables, and evaluated whether markers were shared across - or specific for any domain (**Figure 6; Supplementary Table S4)**. In total, 83 significant omics-disease associations were identified, with 40 metabolites and 17 proteins being significant for ≥1 CIRS domain. Associations were observed with the cardiac, vascular, hypertension, endocrine-metabolic, renal, hepatic, psychiatric, neurological, respiratory and lower gastrointestinal and genitourinary domains (**Figure 6**). Eleven metabolites (serotonin, glutamate, isoleucine, taurine, dihydroxyphenylalanine, several glycerophospholipids and acylcarnitines) and three proteins (F2, C3, A2M) were shared across multiple domains. For example, serotonin was related to the cardiac, vascular, hypertension and psychiatric systems, and glutamate to the hypertension, endocrine-metabolic, respiratory and hepatic domains. The proteins F2 and A2M were related to the cardiac, vascular, and the renal domains, and C3 to the hypertension and endocrine-metabolic domains. Overall, phosphatidylcholines and sphingolipids were all negatively associated to morbidity conditions in the related CIRS domain, whereas for acylcarnitines positive associations were observed. The directionality of biogenic amines and amino acids was not consistent across classes but remained consistent across CIRS domains. For proteins, negative associations were observed with APOB, APOD, APOM, IGHM, CD5L, PON1, FCN3, F2, C4BPA, IGHG2 and IGKC, whereas positive associations were found with SERPINA1, AFM, VTN, C3, HP, SERPIND1 and A2M. In general, all metabolites and proteins that were significantly associated with multiple domains showed consistent effect directions, being either negative or positive.

**Figure 6.**
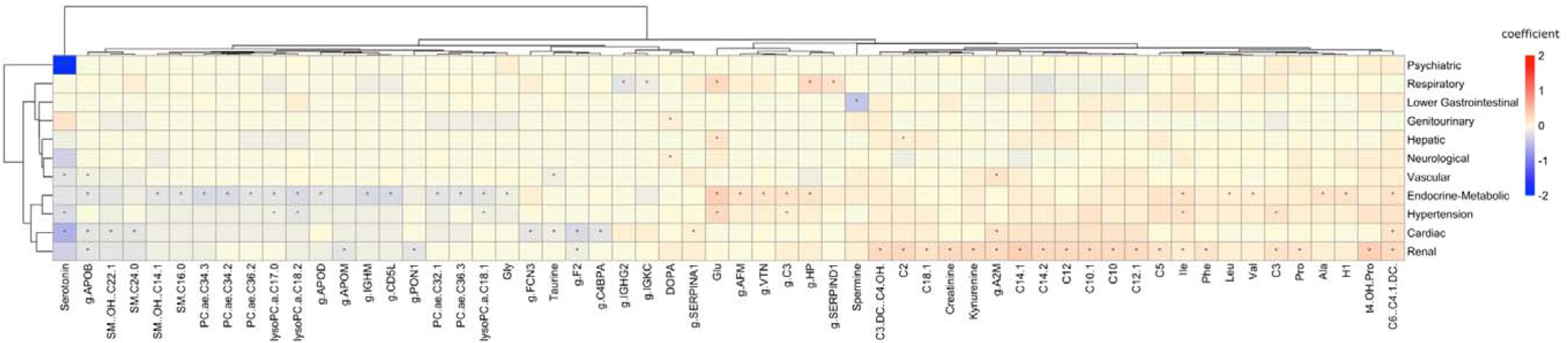
Heatmap presenting associations between metabolites, proteins and CIRS organ domains. Associations were obtained from multiple linear regression models adjusted for age and sex. Hierarchical clustering using the Eucledian distance as the similarity measure was applied. Only metabolites and proteins that were significantly associated with at least one specific CIRS domain are presented. *: highlights significant associations. β-coefficients are color-coded: red hues represent positive relations and blue hues represent negative relations.

## 3. Discussion

In this cross-sectional analysis of CHRIS study data we identified age and 31 molecular features predicting overall health status (any morbidity vs healthy) in adults using a random forest classifier. These features included 21 metabolites (5 biogenic amines; 4 amino acids; 6 acylcarnitines; 5 glycerophospholipids; and hexose monosaccharides), and 10 plasma proteins. Subsequent regression analyses confirmed a sizeable association of health status with both serotonin and glutamate, which was independent of age and sex. Analyses on single CIRS domains further identified multiple metabolite- and protein-disease associations, most being related to cardiovascular, hypertension, endocrine-metabolic and renal morbidity, revealing strong molecular interconnectivity across these related domains.

Several studies have investigated omics markers of aging, longevity^6,19^, aging-related chronic diseases^7,11–13,20,21^ but only few have integrated multi-omics data with regards to general health assessment^15,22^. Independent of the approach used, great heterogeneity exists in the included omics technologies, protein and/or metabolite coverage, analytic tools as well as the outcome of interest, which makes comparison of studies challenging. In our study, multi-omics RF models identified several molecular features relevant for predicting health status (any morbidity vs. healthy).

Among the age-independent predictive features was serotonin, which is involved in the regulation of energy, glucose and lipid metabolism. Changes in the serotonin system are known risk factors for many age-related diseases, such as diabetes and cardiovascular disease^23,24^, which was also observed in our study. Up to 95% of serotonin is produced in the gut, and only 5% of serotonin is synthesized by neurons, mainly in the central nervous system^24^. Although serotonin does not cross the blood-brain barrier, intestinal serotonin release causes neuronal activation in the brain stem, thus indirectly affecting the brain^23^. No other study has linked serotonin as a healthy aging marker per se. However, serotonin is a tryptophan derivative, and as inflammation and stress activate the tryptophan metabolism through the kynurenine pathway^25^, this consequently causes decreased production of serotonin^26^. The age associated upregulation of kynurenine and downregulation of serotonin therefore indicate a relevant role of tryptophan metabolism in inflammaging and aging^27^. Although tryptophan was included in the metabolomics panel in this study, it was not identified among the selected features in the RF model. These results indicate a robust relation between health and circulating serotonin levels, but the role of tryptophan metabolism and related pathways, and the putative causal relationships deserves further investigation.

Glutamate was also identified as an age-independent predictor for health status. This amino acid has been previously associated with physical frailty in elderly individuals^28^, supporting our findings. It has further been linked to cardiovascular disease and been suggested to be a potential biomarker of abdominal obesity and metabolic risk^29^. Glutamate is an important excitatory neurotransmitter in the brain, and although concentrations in the brain are much higher than in plasma as the blood brain barrier is not very permeable to glutamate^30^, it is also one of the most abundant amino acids in the liver, kidney and skeletal muscle, showing great metabolic versatility^31^. Glutamate plays a key role in protein synthesis and degradation^32^ and is a by-product of the catabolism of branched chain amino acids^29^. By linking amino acid and carbohydrate metabolism glutamate supports energy production, which has further implications for insulin secretion^32^. More specifically, glutamate is also a source of alpha-ketoglutarate, which plays a key role in energy metabolism and aging processes and that has been implicated in improved life and health span^33–36^. In this study the morbidity group is associated with higher levels of circulating glutamate, which might reflect a depletion of alpha-ketoglutarate relative to healthy individuals.

Other relevant molecular features (hexose, C18:1, C16:1, lyso PC a C18:2, PC aa C32:1, tyrosine, taurine, kynurenine, AFM, CFH, RBP4, A1BG) were determined with the RF model, and the age independent associations confirmed by linear regression, but the difference in abundances was lower than the cut-off set relative to the coefficient of variation. We provide further discussion of those features in **Supplementary Text S2**.

When looking into organ-specific morbidity, most health conditions in our study were related to the hepatic domain, followed by the vascular, hypertension, endocrine-metabolic and the renal domains. Using OCA, we observed closer relatedness between the renal, hypertension and endocrine-metabolic, as well as the vascular and cardiac domains. Our analyses linking omics markers to specific CIRS domains supported such connections. For example, we observed the metabolites serotonin, glutamate, taurine, isoleucine, three glycerophospholipids (lysoPC a C17.0/18.1/18.2), and two acylcarnitines (C3, C6 (C4)-1-DC) and the proteins C3, APOB, F2, A2M and HP as common markers among the cardiac, vascular, hypertension, endocrine-metabolic, renal but also the respiratory domains. The co-occurrence of type-2 diabetes and cardiovascular diseases is very common, and its high degree of connectivity has been found with other diseases as well^37^. In addition, cardiovascular and kidney disease are closely interrelated and disease of one organ is known to cause dysfunction of the other^38^. Remaining molecular signatures were domain specific showing no connections among each other, such as the proteins VTN, APOD and a set of other amino acids (valine, leucine, alanine) being only related to the endocrine-metabolic domain. Such associations have also been reported previously in the literature^39–41^. Untargeted metabolomics analysis in the prospective, population-based EPIC-Norfolk study identified 420 metabolites that were shared among two or more chronic diseases^12^, further observing high connectivity among cardiometabolic and respiratory diseases across different biochemical classes of metabolites. Those findings highlight potential biological pathways related to the onset of multiple chronic diseases, such as liver and kidney function, lipid and glucose metabolism and low-grade inflammation, among others^12^.

Strengths of our investigation are the wealth of the available data resource, foremost with the availability of both plasma metabolomics and proteomics data among the same participants. Additionally available phenotypic parameters, both quantitative (such as blood parameters), self-reported or collected by trained study-nurses allowed a detailed characterization of the participants and enabled the assessment of the health status through CIRS, which has been shown to be a useful tool to measure morbidity in clinical research^42^.

A first limitations is given by the study design, due to which participants with severe morbidity might have been underrepresented. In addition, metabolite and protein concentrations might be influenced by lifestyle, hormonal changes such as introduced by menopausal status and its treatment, as well as medication. Available information on medication use, irrelevant for the CIRS assessment, allowed adjustments to exclude spurious influence of this factor on the results. However, as the CIRS itself considers medication status to characterize certain diseases, it was not possible to distinguish whether the observed associations were driven by the disease itself or by the treatment for the given or related diseases. The present analyses are also limited to the set of metabolites and proteins that are possible to quantify by the analytical approaches used. Finally, given the cross-sectional design of the study we are only able to assess associations, hence no conclusions on temporal antecedence and causality can be drawn.Finally, we did not validate our models on an independent testing set.Despite these limitations, we were able to replicate several findings from previous studies, which supports reliable data and procedural quality within this study. Overall, using multi-omics data for profiling health status has great potential to identify changes in health trajectories at an earlier stage in life. This could help to develop new, effective target therapies for treating related as well as seemingly unrelated diseases occurring at the same time by uncovering common biological pathways connecting different underlying pathogenic mechanisms^43^.

To conclude, we identified several molecular signatures of overall health status. Specifically, circulating serotonin is suggested as a promising novel predictor for health and morbidity independent of age, implicating a potential key role of tryptophan metabolism and serotonin related pathways in sustaining health. The results also point to glutamate as another predictor for health and morbidity in adults, in agreement with previous studies relating this amino acid to frailty, metabolic and cardiovascular health. Future studies are needed to investigate the mediating role of these signatures in relation to lifestyle and the environment to promote healthy aging. In this regard the application of mendelian randomization approaches should be considered to further investigate causal links between circulating serotonin, serotonin metabolism and chronic disease.

## 4. Methods

### 4.1 Study cohort

The CHRIS study is a population-based cohort of 13,393 adults aged 18 and over recruited from 13 towns in the alpine Val Venosta/Vinschgau district in the Bolzano-South Tyrol province of northern Italy. The study was designed to investigate the genetic and molecular basis of age-related common chronic conditions and their interaction with lifestyle and environment in the general population^44^. The study was approved by the Ethics Committee of the Healthcare System of the Autonomous Province of Bolzano. The study conforms to the Declaration of Helsinki, and with national and institutional legal and ethical requirements.

Metabolomics data were available for n=6,415 individuals and proteomics data for n=3,541. After excluding participants who were not fasting (n=475) or had missing information on fasting status (n=2), as well as women who were pregnant (n=25) or unsure about pregnancy (n=9), n=3,142 individuals with overlapping omics data were available for analysis.

### 4.2 Data collection

Data, including collection of anthropometric measurements, blood and urine standard laboratory tests, blood pressure measurement and lifestyle information were collected at the study center following overnight fasting^44^. Laboratory test data included all main cardiovascular and metabolic risk factors, and markers of iron metabolism, coagulation, renal damage, thyroid, and liver function. Blood (serum and plasma) and urine samples were collected and stored in a biobank.

To obtain information on disease history, participants were interviewed by trained study assistants. Specific clinical domains covered by the questionnaires were the circulatory and nervous system, as well as psychiatric disorders, cognition, autonomic and genitourinary function, endocrine, nutritional, and metabolic diseases. A detailed overview of the mode of assessment can be found elsewhere^44^. In addition, each questionnaire contained a section for “other diseases”, where participants could report any other condition not explicitly included in the domains, as free text. Detailed medication information was collected by scanning the barcodes of the boxes of the medication used within the seven days prior to study center visit and brought by the study participants to the interview. The Anatomical Therapeutic Chemical (ATC) medicinal product classification coding system^45^ and the mode, frequency, and duration of drug administration was recorded for each scanned medication.

### 4.3 Assessment of the health status

To assess health in the CHRIS study sample, we used the Cumulative Illness Rating Scale (CIRS), which is a clinically relevant tool to measure the chronic medical illness burden by taking the number and the severity of chronic diseases into account^46^. For the purpose of this study we used the revised CIRS^18^, which assesses 14 organ related domains rating each of them according to the degree of severity ranging from: grade 0 (no impairment) to grade 4 (extremely severe impairment). Based on this guideline, we determined for all CHRIS participants the CIRS score for each domain by screening our questionnaire and interview data, as well as laboratory and clinical parameters for any given medical condition. We next derived the CIRS Comorbidity Index (CMI), which calculates the total number of CIRS organ domains with a score≥2 (domains with moderate or severe morbidity). To define health status we classified individuals as being healthy if the CMI=0 and as having any morbidity if CMI≥1, which means that at least one CIRS organ domain was identified with moderate or severe morbidity. We additionally considered organ specific health by classifying individuals as being healthy if the CIRS organ domain scored<2, or having morbidity if the respective domain scored≥2.

We further assessed the completeness of the CIRS score with regard to the guidelines^18^ based on available CHRIS data for each domain. Detailed information on the completeness assessment is presented in **Supplementary Text S3**.

#### Metabolite and protein quantification

The AbsoluteIDQ® p180 kit from Biocrates (Biocrates Life Sciences AG, Innsbruck, Austria) was used for metabolite quantification in fasting serum samples. Details on data generation, quality assessment and normalization are provided in Verri Hernandez et al. (2022)^16^. In total, concentrations of 175 metabolites and lipids were quantified. Due to the large number of missing values, sarcosine was excluded from the present analysis. An overview of the included metabolites is presented in the **Supplementary Table S5**. Prior to the main analysis all metabolite concentrations were log2 transformed.

The high abundance plasma proteome was determined using the Scanning SWATH mass spectrometry-based approach. Details on sample processing, data acquisition and normalization are provided in Dordevic et al. (2024)^17^. In total 148 highly abundant proteins were quantified and included in this analysis. An overview of the included proteins is presented in **Supplementary Table S6**.

Metabolite and protein abundances were adjusted for frequent medication use with a linear model based approach: models were fitted separately for each metabolite or protein with their abundance as response, and medication (as individual binary variables) as explanatory variables. Only medications taken at least twice per week and not considered for CIRS scoring were used. The residuals from these models were used to construct medication-independent abundances for the RF and subsequent regression analyses. **Supplementary Table S2** lists the medications for which abundances were adjusted.

### 4.4 Statistical analysis

Differences in characteristics were presented as mean (standard deviation, SD) for continuous variables and as percentages for categorical variables.

To investigate the relations between CIRS-domain specific morbidity (morbidity vs healthy), ordinary correspondence analysis (OCA) was performed using the rda function in the vegan package for R^47^.

#### Random forest

We used a random forest (RF) regression model to predict health status (any morbidity vs healthy), including as predictors age, sex, 174 metabolites and 148 proteins, respectively. Overall, 100 RF models were generated, each containing 500 trees per model, considering 80% training set sizes. Predictions were validated by 100 times stratified repeated random subsampling with 20% test set sizes, such that the ratio between the healthy and the any morbidity group of the original set sizes remained the same for the subsampled sets. We used the randomForest R package^48^ to perform the RF analysis.

Model performance was measured based on the receiver operating characteristic (ROC) area under the curve (AUC), the Matthew’s correlation coefficient (MCC), and the unit-normalized Matthews correlation coefficient (MCC-F1).

The Gini Index was used to measure feature importance. We further estimated the significance of importance metrics for the random forest models by permuting the response variable using the rfpermut*e* R package ^49^. This generates a background distribution and recalculates the 100 random forests, each with 500 trees. Usually, a background distribution is calculated only once; however, to increase the robustness of our results, we created 100 background distributions, yielding 100 *p*-values for each predictor variable. We considered associations with a predictor to be significant if ≥50 out of 100 test runs yielded a *p*-value<0.05.

To further investigate the influence of the significant features obtained from the RF analysis we additionally fitted multiple linear regression models to estimate the association between metabolites and proteins from the selected significant features, each implemented as the response variable, with health status as the explanatory variable and by further adjusting the models for age (years) and sex.

#### Investigation of omics-markers related to CIRS domains

To aid interpretation from the RF analysis, we further investigated the relation between metabolites, proteins and the specific CIRS domains using again multiple linear regression models with implementing all available metabolites and proteins (response variable) one by one with each CIRS domain (explanatory variable), adjusted for age and sex.

We adjusted all *p*-values for multiple hypothesis testing using the Bonferroni correction method (metabolites: 0.05/(14 CIRS domains*174 metabolites); proteins: 0.05/(14 CIRS domains*148 proteins)), thus an adjusted *p*-value <0.05 was considered statistically significant. For all linear regression analyses we additionally required the difference in abundance between categories to be ≥2* coefficient of variation (CV) for the given metabolite and >1*CV for the given protein to consider the markers significant. All analyses were conducted using the R statistical software, version 4.1.0 (www.R-project.org).

## Acknowledgments

CHRIS study investigators thank all study participants, the Healthcare System of the Autonomous Province of Bolzano-South Tyrol, and all Eurac Research staff involved in the study. Bioresource Impact Factor Code: BRIF6107.

## Author contributions

All authors contributed to the study conception and design. Material preparation and data collection were performed by J.R, V.H, F.D, P.P, M.G, L.F, C.P, F.A, V.F, M.R, M.M and E.H. Formal analysis and investigation was performed by C.W, E.H, N.D, J.R and F.D. All authors contributed to the interpretation of the findings. The first draft of the manuscript was written by E.H, C.W, N.D, J.R and F.D and all authors commented on previous versions of the manuscript. All authors read and approved the final manuscript.

## Data availability

The data are not openly available due to reasons of sensitivity. CHRIS study data can be requested for research purposes by submitting a dedicated request to the CHRIS Access Committee.

## Informed Consent Statement

Informed consent was obtained from all subjects involved in the study.

