## Supplementary Information for "Identifying Metabolomic and Proteomic Biomarkers for Age-Related Morbidity in a Population-Based Cohort - the Cooperative Health Research in South Tyrol (CHRIS) study"

**Supplementary Material**

### Supplementary Tables

#### Table S1. Characteristics of the CHRIS cohort with information on health status (any morbidity vs. healthy)^a^ and CIRS domain specific morbidity^b^.

|  | | **Overall** | | **Healthy** | | **Any morbidity** |
| --- | --- | --- | --- | --- | --- | --- |
| Sample size, n | | **13,373^c^** | | **5803** | | **7570** |
| Age, mean (SD) | | 45.3 (16.6) | | 37.9 (13.8) | | 51.0 (16.4) |
| Comorbidity Index, mean (SD) | | 1.02 (1.30) | | 0 | | 1.80 (1.25) |
| Sex n (%) | |  | |  | |  |
| Females | | 7264 (54.3%) | | 3567 (61.5%) | | 3697 (48.8%) |
| Males | | 6109 (45.7%) | | 2236 (38.5%) | | 3873 (51.2%) |
| *Morbidity in CIRS domains*, *yes (%)* | | | | | | |
| Hepatic | 3987 (29.8%) | |  | | 3987 (52.7%) | |
| Vascular | 2024 (15.1%) | |  | | 2024 (26.7%) | |
| Hypertension | 1585 (11.9%) | |  | | 1585 (20.9%) | |
| Respiratory | 1269 (9.5%) | |  | | 1269 (16.8%) | |
| Endocrine-Metabolic | 1422 (10.6%) | |  | | 1422 (18.8%) | |
| Psychiatric and behavioral | 598 (4.5%) | |  | | 598 (7.9%) | |
| Upper Gastrointestinal | 577 (4.3%) | |  | | 577 (7.6%) | |
| MBJ^d^ | 548 (4.1%) | |  | | 548 (7.2%) | |
| Renal | 482 (3.6%) | |  | | 482 (6.4%) | |
| Cardiac | 405 (3.0%) | |  | | 405 (5.4%) | |
| Neurological | 271 (2.0%) | |  | | 271 (3.6%) | |
| Genitourinary | 231 (1.7%) | |  | | 231 (3.1%) | |
| Lower Gastrointestinal | 142 (1.1%) | |  | | 142 (1.9%) | |
| EENT^e^ | 85 (0.6%) | |  | | 85 (1.1%) | |
| ^a^Health status was assessed through the Cumulative Illness Rating Scale (CIRS) Comorbidity Index (CMI) by classifying individuals as having any morbidity (CMI≥1) or being healthy (CMI=0). ^b^CIRS domain specific morbidity is defined as having a score ≥2 in the given domain. ^c^Number of CHRIS baseline study participants available for analysis. ^d^MBJ=Musculoskeletal, bones and joints. ^e^EENT= Ears, eyes, nose and throat. | | | | | | |

#### Table S2. Medications used for adjustment of metabolite and protein abundances.

| **ATC code level 4** | **ATC4 name** | **# participants taking the medication frequently** |
| --- | --- | --- |
| G03AA | Progestogens and estrogens, fixed combinations | 214 |
| A12AX | Calcium, combinations with vitamin D and/or other drugs | 59 |
| G04CA | Alpha-adrenoreceptor antagonists | 40 |
| MO4AA | Preparations inhibiting uric acid production | 31 |
| G03AB | Progestogens and estrogens, sequential preparations | 27 |
| G02BA | Intrauterine contraceptives | 22 |
| S01ED | Beta blocking agents | 18 |
| G03HB | Antiandrogens and estrogens | 14 |
| N04BC | Dopamine agonists | 14 |
| G03FA | Progestogens and estrogens, fixed combinations | 13 |
| G03CA | Natural and semisynthetic estrogens, plain | 10 |
| G03AC | Progestogens | 8 |
| J01FA | Macrolides | 8 |
| A11DB | Vitamin B1 in combination with vitamin B6 and/or vitamin B12 | 7 |
| G04CB | Testosterone-5-alpha reductase inhibitors | 7 |
| R01AD | Corticosteroids | 6 |
| R06AE | Piperazine derivatives | 6 |
| R06AX | Other antihistamines for systemic use | 6 |
| S01EE | Prostaglandin analogues | 6 |

Table S3. Results of the regression analyses for association between health status, age and metabolite or protein abundances for significant features obtained from the random forest analysis.

| **Feature name** | **Description** | **Class/Protein ID** | **coef-anymorb** | **%diff-anymorb** | **p-adj-anymorb** | **coef-age** | **p-adj-age** | **CV QC (%)** |
| --- | --- | --- | --- | --- | --- | --- | --- | --- |
| *Metabolites* | | | | | | | | |
| Serotonin | Serotonin | biogenic amines | -0.287 | 22.031 | 1.03E-16 | -0.006 | 4.52E-09 | 3.926 |
| Glu | Glutamate | amino acids | 0.160 | 11.754 | 8.27E-13 | 0.004 | 1.12E-09 | 3.756 |
| Tyr | Tyrosine | amino acids | 0.063 | 4.451 | 1.55E-06 | 0.001 | 4.62E-04 | 3.808 |
| H1 | Hexose | sugars | 0.034 | 2.404 | 3.34E-04 | 0.003 | 1.65E-27 | 3.299 |
| PC aa C32:1 | Phosphatidylcholine diacyl C32:1 | glycerophospholipids | 0.118 | 8.530 | 3.80E-04 | 0.005 | 8.42E-10 | 7.670 |
| lysoPC a C18:2 | lysoPhosphatidylcholine acyl C18:2 | glycerophospholipids | -0.079 | 5.645 | 4.23E-04 | -0.004 | 5.11E-10 | 4.475 |
| C18:1 | Octadecenoylcarnitine | acylcarnitines | 0.056 | 3.953 | 6.23E-04 | 0.004 | 2.98E-20 | 5.348 |
| Kynurenine | Kynurenine | biogenic amines | 0.049 | 3.427 | 5.66E-03 | 0.002 | 1.41E-08 | 6.675 |
| C16:1 | Hexadecenoylcarnitine | acylcarnitines | 0.045 | 3.149 | 8.88E-03 | 0.004 | 3.11E-30 | 7.481 |
| Taurine | Taurine | biogenic amines | -0.040 | 2.777 | 1.13E-02 | 0.000 | 1 | 3.036 |
| C6 (C4:1-DC) | Hexanoylcarnitine (Fumarylcarnitine) | acylcarnitines | 0.049 | 3.419 | 8.03E-02 | 0.006 | 4.57E-29 | 6.375 |
| C16 | Hexadecanoylcarnitine | acylcarnitines | 0.038 | 2.698 | 1.71E-01 | 0.004 | 1.51E-18 | 18.773 |
| PC aa C40:4 | Phosphatidylcholine diacyl C40:4 | glycerophospholipids | 0.047 | 3.305 | 3.33E-01 | 0.004 | 1.24E-10 | 7.155 |
| PC aa C38:4 | Phosphatidylcholine diacyl C38:4 | glycerophospholipids | 0.044 | 3.094 | 4.38E-01 | 0.004 | 5.19E-14 | 5.153 |
| C0 | Carnitine | acylcarnitines | 0.027 | 1.860 | 5.60E-01 | 0.002 | 3.81E-08 | 5.650 |
| PC aa C40:5 | Phosphatidylcholine diacyl C40:5 | glycerophospholipids | 0.041 | 2.848 | 8.47E-01 | 0.006 | 2.13E-26 | 6.630 |
| alpha-AAA | alpha-Aminoadipic acid | biogenic amines | -0.038 | 2.669 | 1 | 0.006 | 2.05E-02 | 21.487 |
| Cit | Citrulline | amino acids | -0.022 | 1.522 | 1 | 0.007 | 2.99E-67 | 4.038 |
| Orn | Ornithine | amino acids | 0.035 | 2.429 | 1 | 0.004 | 1.01E-10 | 6.629 |
| SDMA | Symmetric dimethylarginine | biogenic amines | -0.005 | 0.322 | 1 | 0.004 | 7.55E-36 | 8.146 |
| C18 | Octadecanoylcarnitine | acylcarnitines | 0.000 | 0.016 | 1 | 0.005 | 5.55E-30 | 16.274 |
| *Proteins* | | | | | | | | |
| g:AFM | Afamin | P43652 | 0.071 | 5.075 | 2.23E-08 | -0.001 | 2.11E-02 | 9.574 |
| g:A1BG* | Alpha-1B-glycoprotein | P04217 | 0.036 | 2.550 | 2.45E-04 | 0.002 | 4.36E-15 | 9.357 |
| g:CFH | Complement factor H | P08603 | 0.035 | 2.447 | 4.98E-04 | 0.002 | 2.49E-08 | 7.930 |
| g:RBP4 | Retinol-binding protein 4 | P02753 | 0.043 | 3.034 | 3.36E-03 | -0.001 | 1.01E-01 | 9.483 |
| g:A2M | Alpha-2-macroglobulin | P01023 | -0.041 | 2.897 | 8.88E-02 | -0.002 | 1.18E-05 | 7.058 |
| g:APOH | Beta-2-glycoprotein 1 | P02749 | 0.034 | 2.382 | 3.36E-01 | 0.002 | 5.00E-05 | 18.547 |
| g:F2 | Prothrombin | P00734 | -0.016 | 1.116 | 3.57E-01 | -0.002 | 5.37E-20 | 7.021 |
| g:IGHM | Immunoglobulin heavy constant mu | P01871 | -0.048 | 3.377 | 9.72E-01 | -0.007 | 1.67E-14 | 10.951 |
| g:IGFALS | Insulin-like growth factor-binding protein complex acid labile subunit | P35858;P35858-2 | -0.002 | 0.148 | 1 | -0.009 | 2.48E-81 | 20.125 |
| g:C4BPA | C4b-binding protein alpha chain | P04003 | 0.014 | 0.972 | 1 | 0.004 | 2.59E-27 | 8.639 |
| Features are sorted by increasing *p*-value for the trait health status (any morbidity vs. healthy). *: Metabolites and proteins with a Bonferroni adjusted *p*-value<0.05 and a difference in concentrations (expressed in %) larger than 2 times or one time the coefficient of variation (CV) in QC (study pool) samples (expressed in %) for metabolites and proteins, respectively, are considered statistically significant. coef-anymorb: coefficient for the trait any morbidity vs. healthy representing the average differential abundance in log2 scale between the compared groups. Positive values indicate higher, negative values lower, average abundances in individuals with any morbidity compared to those without. coef-age: coefficient for the trait age representing the change in log2 abundance in one year. Positive values indicate increasing, negative values decreasing abundances with age. %difference: represents the difference of (mean) concentrations on normal scale in %. *p*-adj-anymorb: *p*-value for the trait any morbidity vs healthy, adjusted for multiple hypothesis testing using the Bonferroni method. *p*-adj-age *p*-value for the trait age, adjusted for multiple hypothesis testing using the Bonferroni method. | | | | | | | | |

Table S4. Results of the regression analyses for significant association between morbidity in CIRS organ domains and metabolite or protein abundances.

| **Feature** | **Description** | **Class/Protein ID** | **CIRS domain** | **Coef** | **p-adj** | **%diff** | **CV QC** |
| --- | --- | --- | --- | --- | --- | --- | --- |
| *Metabolites* |  |  |  |  |  |  |  |
| Serotonin | Serotonin | biogenic amines | Cardiac | -0.588 | 4.57E-09 | 50.276 | 3.926 |
| Taurine | Taurine | biogenic amines | Cardiac | -0.126 | 2.55E-02 | 9.140 | 3.036 |
| C6 (C4:1-DC) | Hexanoylcarnitine (Fumarylcarnitine) | acylcarnitines | Cardiac | 0.188 | 1.82E-02 | 13.939 | 6.375 |
| SM (OH) C22:1 | Hydroxysphingomyeline C22:1 | sphingolipids | Cardiac | -0.217 | 2.17E-03 | 16.195 | 7.374 |
| SM C24:0 | Sphingomyeline C24:0 | sphingolipids | Cardiac | -0.216 | 8.75E-04 | 16.141 | 6.872 |
| Ala | Alanine | amino acids | Endocrine-Metabolic | 0.145 | 4.79E-12 | 10.582 | 4.790 |
| Glu | Glutamate | amino acids | Endocrine-Metabolic | 0.355 | 9.92E-24 | 27.933 | 3.756 |
| Gly | Glycine | amino acids | Endocrine-Metabolic | -0.121 | 6.47E-05 | 8.756 | 3.743 |
| Ile | Isoleucine | amino acids | Endocrine-Metabolic | 0.162 | 4.26E-19 | 11.855 | 3.691 |
| Leu | Leucine | amino acids | Endocrine-Metabolic | 0.114 | 5.03E-10 | 8.199 | 4.051 |
| Val | Valine | amino acids | Endocrine-Metabolic | 0.115 | 3.05E-10 | 8.281 | 4.040 |
| C6 (C4:1-DC) | Hexanoylcarnitine (Fumarylcarnitine) | acylcarnitines | Endocrine-Metabolic | 0.190 | 7.95E-10 | 14.082 | 6.375 |
| H1 | Hexose | sugars | Endocrine-Metabolic | 0.171 | 8.00E-41 | 12.553 | 3.299 |
| lysoPC a C17:0 | lysoPhosphatidylcholine acyl C17:0 | glycerophospholipids | Endocrine-Metabolic | -0.222 | 1.64E-13 | 16.645 | 5.380 |
| lysoPC a C18:1 | lysoPhosphatidylcholine acyl C18:1 | glycerophospholipids | Endocrine-Metabolic | -0.168 | 1.66E-07 | 12.331 | 3.155 |
| lysoPC a C18:2 | lysoPhosphatidylcholine acyl C18:2 | glycerophospholipids | Endocrine-Metabolic | -0.248 | 2.43E-14 | 18.737 | 4.475 |
| PC ae C32:1 | Phosphatidylcholine acyl-alkyl C32:1 | glycerophospholipids | Endocrine-Metabolic | -0.198 | 5.37E-10 | 14.676 | 7.084 |
| PC ae C34:2 | Phosphatidylcholine acyl-alkyl C34:2 | glycerophospholipids | Endocrine-Metabolic | -0.264 | 9.17E-15 | 20.050 | 6.374 |
| PC ae C34:3 | Phosphatidylcholine acyl-alkyl C34:3 | glycerophospholipids | Endocrine-Metabolic | -0.292 | 3.20E-16 | 22.430 | 6.138 |
| PC ae C36:2 | Phosphatidylcholine acyl-alkyl C36:2 | glycerophospholipids | Endocrine-Metabolic | -0.228 | 7.26E-12 | 17.124 | 6.473 |
| PC ae C36:3 | Phosphatidylcholine acyl-alkyl C36:3 | glycerophospholipids | Endocrine-Metabolic | -0.206 | 4.27E-09 | 15.332 | 6.131 |
| SM (OH) C14:1 | Hydroxysphingomyeline C14:1 | sphingolipids | Endocrine-Metabolic | -0.188 | 1.18E-07 | 13.911 | 6.696 |
| SM C16:0 | Sphingomyeline C16:0 | sphingolipids | Endocrine-Metabolic | -0.174 | 3.59E-09 | 12.842 | 5.838 |
| DOPA | Dihydroxyphenylalanine | biogenic amines | Genitourinary | 0.104 | 1.32E-02 | 7.504 | 2.927 |
| Glu | Glutamate | amino acids | Hepatic | 0.127 | 5.34E-05 | 9.211 | 3.756 |
| C2 | Acetylcarnitine | acylcarnitines | Hepatic | 0.104 | 2.78E-04 | 7.451 | 2.333 |
| Glu | Glutamate | amino acids | Hypertension | 0.207 | 1.41E-07 | 15.408 | 3.756 |
| Ile | Isoleucine | amino acids | Hypertension | 0.124 | 1.27E-11 | 8.956 | 3.691 |
| Serotonin | Serotonin | biogenic amines | Hypertension | -0.290 | 1.09E-05 | 22.303 | 3.926 |
| C3 | Propionylcarnitine | acylcarnitines | Hypertension | 0.154 | 6.14E-07 | 11.256 | 4.274 |
| lysoPC a C17:0 | lysoPhosphatidylcholine acyl C17:0 | glycerophospholipids | Hypertension | -0.158 | 1.21E-06 | 11.569 | 5.380 |
| lysoPC a C18:1 | lysoPhosphatidylcholine acyl C18:1 | glycerophospholipids | Hypertension | -0.124 | 1.05E-03 | 8.969 | 3.155 |
| lysoPC a C18:2 | lysoPhosphatidylcholine acyl C18:2 | glycerophospholipids | Hypertension | -0.166 | 4.71E-06 | 12.172 | 4.475 |
| Spermine | Spermine | biogenic amines | Lower_GI | -0.420 | 6.53E-04 | 33.770 | 15.843 |
| DOPA | Dihydroxyphenylalanine | biogenic amines | Neurological | 0.093 | 3.14E-03 | 6.649 | 2.927 |
| Serotonin | Serotonin | biogenic amines | Psychiatric | -2.203 | 1.87E-245 | 360.496 | 3.926 |
| Creatinine | Creatinine | biogenic amines | Renal | 0.227 | 2.31E-18 | 17.011 | 4.715 |
| Ile | Isoleucine | amino acids | Renal | 0.121 | 3.57E-03 | 8.767 | 3.691 |
| Kynurenine | Kynurenine | biogenic amines | Renal | 0.253 | 1.90E-12 | 19.138 | 6.675 |
| Phe | Phenylalanine | amino acids | Renal | 0.131 | 1.50E-05 | 9.478 | 3.829 |
| Pro | Proline | amino acids | Renal | 0.160 | 3.05E-02 | 11.702 | 5.280 |
| t4-OH-Pro | trans-4-Hydroxyproline | biogenic amines | Renal | 0.309 | 1.98E-06 | 23.916 | 7.068 |
| C10 | Decanoylcarnitine | acylcarnitines | Renal | 0.228 | 9.91E-03 | 17.131 | 4.140 |
| C10:1 | Decenoylcarnitine | acylcarnitines | Renal | 0.252 | 2.66E-07 | 19.106 | 5.294 |
| C12 | Dodecanoylcarnitine | acylcarnitines | Renal | 0.223 | 1.46E-05 | 16.740 | 7.442 |
| C12:1 | Dodecenoylcarnitine | acylcarnitines | Renal | 0.270 | 2.67E-09 | 20.586 | 5.369 |
| C14:1 | Tetradecenoylcarnitine | acylcarnitines | Renal | 0.298 | 4.29E-05 | 22.975 | 5.826 |
| C14:2 | Tetradecadienylcarnitine | acylcarnitines | Renal | 0.222 | 9.21E-03 | 16.675 | 7.463 |
| C18:1 | Octadecenoylcarnitine | acylcarnitines | Renal | 0.147 | 8.23E-03 | 10.709 | 5.348 |
| C2 | Acetylcarnitine | acylcarnitines | Renal | 0.218 | 1.04E-03 | 16.328 | 2.333 |
| C3 | Propionylcarnitine | acylcarnitines | Renal | 0.176 | 1.22E-02 | 13.007 | 4.274 |
| C3-DC (C4-OH) | Hydroxybutyrylcarnitine | acylcarnitines | Renal | 0.229 | 5.18E-04 | 17.202 | 7.986 |
| C5 | Valerylcarnitine | acylcarnitines | Renal | 0.182 | 3.67E-03 | 13.418 | 5.324 |
| C6 (C4:1-DC) | Hexanoylcarnitine (Fumarylcarnitine) | acylcarnitines | Renal | 0.252 | 4.30E-07 | 19.071 | 6.375 |
| Glu | Glutamate | amino acids | Respiratory | 0.233 | 1.72E-08 | 17.554 | 3.756 |
| Serotonin | Serotonin | biogenic amines | Vascular | -0.228 | 6.66E-04 | 17.115 | 3.926 |
| Taurine | Taurine | biogenic amines | Vascular | -0.120 | 5.21E-12 | 8.679 | 3.036 |
| *Proteins* |  |  |  |  |  |  |  |
| g:F2 | Prothrombin | P00734 | Cardiac | -0.246 | 2.51E-35 | 18.552 | 7.021 |
| g:FCN3 | Ficolin-3 | O75636 | Cardiac | -0.240 | 3.11E-02 | 18.063 | 17.674 |
| g:C4BPA | C4b-binding protein alpha chain | P04003 | Cardiac | -0.228 | 4.17E-14 | 17.141 | 8.639 |
| g:APOB | Apolipoprotein B-100 | P04114 | Cardiac | -0.227 | 9.79E-06 | 17.014 | 9.899 |
| g:A2M | Alpha-2-macroglobulin | P01023 | Cardiac | 0.189 | 3.59E-03 | 13.971 | 7.058 |
| g:SERPINA1 | Alpha-1-antitrypsin | P01009 | Cardiac | 0.120 | 2.22E-02 | 8.669 | 7.950 |
| g:IGHM | Immunoglobulin heavy constant mu | P01871 | Endocrine-Metabolic | -0.269 | 5.05E-06 | 20.458 | 10.951 |
| g:CD5L | CD5 antigen-like | O43866 | Endocrine-Metabolic | -0.249 | 4.64E-05 | 18.855 | 17.787 |
| g:APOD | Apolipoprotein D | P05090 | Endocrine-Metabolic | -0.223 | 2.78E-17 | 16.700 | 13.115 |
| g:APOB | Apolipoprotein B-100 | P04114 | Endocrine-Metabolic | -0.182 | 7.24E-11 | 13.468 | 9.899 |
| g:HP | Haptoglobin | P00738 | Endocrine-Metabolic | 0.180 | 3.69E-03 | 13.259 | 10.070 |
| g:VTN | Vitronectin | P04004 | Endocrine-Metabolic | 0.135 | 2.44E-20 | 9.794 | 8.553 |
| g:AFM | Afamin | P43652 | Endocrine-Metabolic | 0.134 | 1.25E-09 | 9.743 | 9.575 |
| g:C3 | Complement C3 | P01024 | Endocrine-Metabolic | 0.131 | 2.24E-24 | 9.511 | 4.871 |
| g:C3 | Complement C3 | P01024 | Hypertension | 0.102 | 1.90E-15 | 7.294 | 4.871 |
| g:A2M | Alpha-2-macroglobulin | P01023 | Renal | 0.275 | 2.04E-10 | 21.037 | 7.058 |
| g:APOM | Apolipoprotein M | O95445 | Renal | -0.214 | 4.76E-08 | 15.988 | 14.761 |
| g:APOB | Apolipoprotein B-100 | P04114 | Renal | -0.169 | 7.32E-03 | 12.427 | 9.899 |
| g:PON1 | Serum paraoxonase/arylesterase 1 | P27169 | Renal | -0.169 | 3.49E-02 | 12.418 | 11.318 |
| g:F2 | Prothrombin | P00734 | Renal | -0.130 | 8.04E-10 | 9.440 | 7.021 |
| g:HP | Haptoglobin | P00738 | Respiratory | 0.237 | 1.71E-06 | 17.842 | 10.070 |
| g:IGHG2 | Immunoglobulin heavy constant gamma 2 | P01859 | Respiratory | -0.212 | 2.15E-09 | 15.801 | 8.071 |
| g:SERPIND1 | Heparin cofactor 2 | P05546 | Respiratory | 0.149 | 2.85E-09 | 10.917 | 7.994 |
| g:IGKC | Immunoglobulin kappa constant | P01834 | Respiratory | -0.120 | 4.76E-06 | 8.666 | 7.620 |
| g:APOB | Apolipoprotein B-100 | P04114 | Vascular | -0.147 | 1.48E-09 | 10.738 | 9.899 |
| g:A2M | Alpha-2-macroglobulin | P01023 | Vascular | 0.142 | 1.93E-08 | 10.373 | 7.058 |
| Metabolites and proteins related to specific CIRS domains with a Bonferroni adjusted p-value<0.05 and a difference in concentrations (expressed in %) larger than 2 times or one time the coefficient of variation (CV) in QC (study pool) samples (expressed in %) for metabolites and proteins, respectively, are considered statistically significant. coef: coefficient representing the average differential abundance in log2 scale between the compared groups (morbidity in CIRS domain vs. healthy). Positive values indicate higher, negative values lower, average abundances in individuals with CIRS domain specific morbidity compared to those without. %diff: represents the difference of (mean) concentrations on normal scale in %. p-adj: p-value adjusted for multiple hypothesis testing using the Bonferroni method. | | | | | | | |

Table S5. Overview of the 174 metabolites included in the analysis. 14 out of the in total 188 metabolites quantified with the Biocrates AbsoluteIDQ p180 kit were excluded because of poor quality or high number of missing values in the CHRIS cohort.

| **Analyte name** | **Biochemical name** | **Analyte class** | **Mean** | **Sd** | **CV QC (%)** |
| --- | --- | --- | --- | --- | --- |
| ADMA | Asymmetric dimethylarginine | biogenic amines | 0.49 | 3.20 | 6.63 |
| Ala | Alanine | amino acids | 344.80 | 72.83 | 4.79 |
| alpha-AAA | alpha-Aminoadipic acid | biogenic amines | 0.57 | 0.48 | 21.49 |
| Arg | Arginine | amino acids | 101.52 | 18.57 | 3.82 |
| Asn | Asparagine | amino acids | 45.74 | 7.28 | 3.14 |
| Asp | Aspartate | amino acids | 14.73 | 3.98 | 5.46 |
| Cit | Citrulline | amino acids | 30.05 | 7.90 | 4.04 |
| Creatinine | Creatinine | biogenic amines | 77.28 | 19.61 | 4.71 |
| DOPA | Dihydroxyphenylalanine | biogenic amines | 0.19 | 0.11 | 2.93 |
| Gln | Glutamine | amino acids | 646.42 | 97.92 | 4.85 |
| Glu | Glutamate | amino acids | 46.15 | 19.15 | 3.76 |
| Gly | Glycine | amino acids | 248.73 | 72.65 | 3.74 |
| His | Histidine | amino acids | 90.08 | 12.06 | 3.54 |
| Histamine | Histamine | biogenic amines | 0.14 | 0.02 | 0.99 |
| Ile | Isoleucine | amino acids | 66.02 | 15.21 | 3.69 |
| Kynurenine | Kynurenine | biogenic amines | 2.70 | 0.73 | 6.68 |
| Leu | Leucine | amino acids | 132.71 | 27.36 | 4.05 |
| Lys | Lysine | amino acids | 207.00 | 39.12 | 7.07 |
| Met | Methionine | amino acids | 22.78 | 4.26 | 4.85 |
| Met-SO | Methioninesulfoxide | biogenic amines | 0.23 | 0.37 | 51.51 |
| Orn | Ornithine | amino acids | 73.00 | 19.34 | 6.63 |
| Phe | Phenylalanine | amino acids | 63.71 | 9.63 | 3.83 |
| Pro | Proline | amino acids | 174.01 | 56.44 | 5.28 |
| Putrescine | Putrescine | biogenic amines | 0.14 | 0.05 | 8.71 |
| SDMA | Symmetric dimethylarginine | biogenic amines | 0.51 | 2.79 | 8.15 |
| Ser | Serine | amino acids | 124.46 | 21.76 | 4.49 |
| Serotonin | Serotonin | biogenic amines | 0.69 | 0.32 | 3.93 |
| Spermidine | Spermidine | biogenic amines | 0.20 | 0.04 | 5.21 |
| Spermine | Spermine | biogenic amines | 0.16 | 0.03 | 15.84 |
| t4-OH-Pro | trans-4-Hydroxyproline | biogenic amines | 8.60 | 6.07 | 7.07 |
| Taurine | Taurine | biogenic amines | 108.26 | 21.31 | 3.04 |
| Thr | Threonine | amino acids | 121.16 | 26.98 | 3.44 |
| Trp | Tryptophan | amino acids | 61.90 | 10.45 | 3.55 |
| Tyr | Tyrosine | amino acids | 64.71 | 14.19 | 3.81 |
| Val | Valine | amino acids | 220.60 | 43.32 | 4.04 |
| C0 | Carnitine | acylcarnitines | 38.45 | 8.91 | 5.65 |
| C10 | Decanoylcarnitine | acylcarnitines | 0.36 | 0.18 | 4.14 |
| C10:1 | Decenoylcarnitine | acylcarnitines | 0.16 | 0.05 | 5.29 |
| C10:2 | Decadienylcarnitine | acylcarnitines | 0.08 | 0.01 | 9.16 |
| C12 | Dodecanoylcarnitine | acylcarnitines | 0.14 | 0.05 | 7.44 |
| C12-DC | Dodecanedioylcarnitine | acylcarnitines | 0.33 | 0.03 | 4.57 |
| C12:1 | Dodecenoylcarnitine | acylcarnitines | 0.15 | 0.04 | 5.37 |
| C14 | Tetradecanoylcarnitine | acylcarnitines | 0.06 | 0.02 | 16.24 |
| C14:1 | Tetradecenoylcarnitine | acylcarnitines | 0.08 | 0.04 | 5.83 |
| C14:1-OH | Hydroxytetradecenoylcarnitine | acylcarnitines | 0.03 | 0.01 | 9.12 |
| C14:2 | Tetradecadienylcarnitine | acylcarnitines | 0.04 | 0.02 | 7.46 |
| C14:2-OH | Hydroxytetradecadienylcarnitine | acylcarnitines | 0.03 | 0.01 | 8.47 |
| C16 | Hexadecanoylcarnitine | acylcarnitines | 0.13 | 0.03 | 18.77 |
| C16-OH | Hydroxyhexadecanoylcarnitine | acylcarnitines | 0.03 | 0.01 | 8.81 |
| C16:1 | Hexadecenoylcarnitine | acylcarnitines | 0.05 | 0.01 | 7.48 |
| C16:1-OH | Hydroxyhexadecenoylcarnitine | acylcarnitines | 0.02 | 0.00 | 12.17 |
| C18 | Octadecanoylcarnitine | acylcarnitines | 0.07 | 0.02 | 16.27 |
| C18:1 | Octadecenoylcarnitine | acylcarnitines | 0.13 | 0.03 | 5.35 |
| C2 | Acetylcarnitine | acylcarnitines | 7.61 | 2.68 | 2.33 |
| C3 | Propionylcarnitine | acylcarnitines | 0.38 | 0.13 | 4.27 |
| C3-DC (C4-OH) | Hydroxybutyrylcarnitine | acylcarnitines | 0.05 | 0.02 | 7.99 |
| C3:1 | Propenoylcarnitine | acylcarnitines | 0.01 | 0.00 | 14.31 |
| C4 | Butyrylcarnitine | acylcarnitines | 0.21 | 0.10 | 6.25 |
| C4:1 | Butenylcarnitine | acylcarnitines | 0.02 | 0.00 | 11.91 |
| C5 | Valerylcarnitine | acylcarnitines | 0.13 | 0.05 | 5.32 |
| C5-DC (C6-OH) | Glutarylcarnitine (Hydroxyhexanoylcarnitine) | acylcarnitines | 0.02 | 0.01 | 11.56 |
| C5-M-DC | Methylglutarylcarnitine | acylcarnitines | 0.03 | 0.01 | 11.13 |
| C5-OH (C3-DC-M) | Hydroxyvalerylcarnitine (Methylmalonylcarnitine) | acylcarnitines | 0.05 | 0.01 | 8.71 |
| C5:1 | Tiglylcarnitine | acylcarnitines | 0.03 | 0.01 | 9.65 |
| C5:1-DC | Glutaconylcarnitine | acylcarnitines | 0.02 | 0.00 | 12.52 |
| C6 (C4:1-DC) | Hexanoylcarnitine (Fumarylcarnitine) | acylcarnitines | 0.07 | 0.03 | 6.38 |
| C6:1 | Hexenoylcarnitine | acylcarnitines | 0.02 | 0.00 | 11.21 |
| C8 | Octanoylcarnitine | acylcarnitines | 0.22 | 0.11 | 8.83 |
| C9 | Nonaylcarnitine | acylcarnitines | 0.07 | 0.02 | 7.42 |
| H1 | Hexose | sugars | 4491.72 | 750.67 | 3.30 |
| lysoPC a C14:0 | lysoPhosphatidylcholine acyl C14:0 | glycerophospholipids | 3.63 | 0.50 | 6.03 |
| lysoPC a C16:0 | lysoPhosphatidylcholine acyl C16:0 | glycerophospholipids | 58.18 | 12.81 | 4.56 |
| lysoPC a C16:1 | lysoPhosphatidylcholine acyl C16:1 | glycerophospholipids | 2.13 | 0.69 | 7.00 |
| lysoPC a C17:0 | lysoPhosphatidylcholine acyl C17:0 | glycerophospholipids | 1.27 | 0.39 | 5.38 |
| lysoPC a C18:0 | lysoPhosphatidylcholine acyl C18:0 | glycerophospholipids | 18.59 | 5.15 | 6.48 |
| lysoPC a C18:1 | lysoPhosphatidylcholine acyl C18:1 | glycerophospholipids | 14.79 | 4.70 | 3.15 |
| lysoPC a C18:2 | lysoPhosphatidylcholine acyl C18:2 | glycerophospholipids | 22.06 | 8.14 | 4.47 |
| lysoPC a C20:3 | lysoPhosphatidylcholine acyl C20:3 | glycerophospholipids | 2.09 | 0.67 | 4.88 |
| lysoPC a C20:4 | lysoPhosphatidylcholine acyl C20:4 | glycerophospholipids | 5.37 | 1.79 | 4.52 |
| lysoPC a C24:0 | lysoPhosphatidylcholine acyl C24:0 | glycerophospholipids | 0.10 | 0.02 | 15.55 |
| lysoPC a C26:0 | lysoPhosphatidylcholine acyl C26:0 | glycerophospholipids | 0.10 | 0.03 | 18.36 |
| lysoPC a C26:1 | lysoPhosphatidylcholine acyl C26:1 | glycerophospholipids | 0.06 | 0.02 | 18.74 |
| lysoPC a C28:0 | lysoPhosphatidylcholine acyl C28:0 | glycerophospholipids | 0.17 | 0.04 | 13.62 |
| lysoPC a C28:1 | lysoPhosphatidylcholine acyl C28:1 | glycerophospholipids | 0.26 | 0.08 | 10.36 |
| PC aa C24:0 | Phosphatidylcholine diacyl C24:0 | glycerophospholipids | 0.06 | 0.09 | 257.42 |
| PC aa C26:0 | Phosphatidylcholine diacyl C26:0 | glycerophospholipids | 0.49 | 0.07 | 7.58 |
| PC aa C28:1 | Phosphatidylcholine diacyl C28:1 | glycerophospholipids | 2.93 | 0.86 | 6.20 |
| PC aa C30:0 | Phosphatidylcholine diacyl C30:0 | glycerophospholipids | 3.49 | 1.28 | 6.32 |
| PC aa C30:2 | Phosphatidylcholine diacyl C30:2 | glycerophospholipids | 0.21 | 0.11 | 58.25 |
| PC aa C32:0 | Phosphatidylcholine diacyl C32:0 | glycerophospholipids | 35.66 | 8.61 | 6.54 |
| PC aa C32:1 | Phosphatidylcholine diacyl C32:1 | glycerophospholipids | 51.14 | 27.80 | 7.67 |
| PC aa C32:2 | Phosphatidylcholine diacyl C32:2 | glycerophospholipids | 15.03 | 6.17 | 8.51 |
| PC aa C32:3 | Phosphatidylcholine diacyl C32:3 | glycerophospholipids | 1.43 | 0.43 | 8.42 |
| PC aa C34:1 | Phosphatidylcholine diacyl C34:1 | glycerophospholipids | 479.48 | 148.61 | 6.77 |
| PC aa C34:2 | Phosphatidylcholine diacyl C34:2 | glycerophospholipids | 1095.60 | 278.45 | 5.98 |
| PC aa C34:3 | Phosphatidylcholine diacyl C34:3 | glycerophospholipids | 37.45 | 12.94 | 5.82 |
| PC aa C34:4 | Phosphatidylcholine diacyl C34:4 | glycerophospholipids | 5.30 | 2.11 | 6.77 |
| PC aa C36:0 | Phosphatidylcholine diacyl C36:0 | glycerophospholipids | 3.97 | 1.27 | 11.85 |
| PC aa C36:1 | Phosphatidylcholine diacyl C36:1 | glycerophospholipids | 64.37 | 20.03 | 7.19 |
| PC aa C36:2 | Phosphatidylcholine diacyl C36:2 | glycerophospholipids | 424.21 | 107.47 | 5.72 |
| PC aa C36:3 | Phosphatidylcholine diacyl C36:3 | glycerophospholipids | 256.96 | 71.65 | 6.15 |
| PC aa C36:4 | Phosphatidylcholine diacyl C36:4 | glycerophospholipids | 389.24 | 114.39 | 5.39 |
| PC aa C36:5 | Phosphatidylcholine diacyl C36:5 | glycerophospholipids | 39.48 | 19.22 | 6.25 |
| PC aa C36:6 | Phosphatidylcholine diacyl C36:6 | glycerophospholipids | 1.75 | 0.73 | 8.51 |
| PC aa C38:0 | Phosphatidylcholine diacyl C38:0 | glycerophospholipids | 3.33 | 1.01 | 6.50 |
| PC aa C38:1 | Phosphatidylcholine diacyl C38:1 | glycerophospholipids | 1.18 | 0.66 | 44.81 |
| PC aa C38:3 | Phosphatidylcholine diacyl C38:3 | glycerophospholipids | 65.03 | 20.68 | 5.85 |
| PC aa C38:4 | Phosphatidylcholine diacyl C38:4 | glycerophospholipids | 153.27 | 45.87 | 5.15 |
| PC aa C38:5 | Phosphatidylcholine diacyl C38:5 | glycerophospholipids | 76.58 | 21.51 | 5.09 |
| PC aa C38:6 | Phosphatidylcholine diacyl C38:6 | glycerophospholipids | 104.84 | 35.65 | 5.92 |
| PC aa C40:1 | Phosphatidylcholine diacyl C40:1 | glycerophospholipids | 0.40 | 0.09 | 11.68 |
| PC aa C40:2 | Phosphatidylcholine diacyl C40:2 | glycerophospholipids | 0.30 | 0.09 | 15.82 |
| PC aa C40:3 | Phosphatidylcholine diacyl C40:3 | glycerophospholipids | 0.60 | 0.15 | 11.62 |
| PC aa C40:4 | Phosphatidylcholine diacyl C40:4 | glycerophospholipids | 3.75 | 1.19 | 7.15 |
| PC aa C40:5 | Phosphatidylcholine diacyl C40:5 | glycerophospholipids | 9.81 | 3.18 | 6.63 |
| PC aa C40:6 | Phosphatidylcholine diacyl C40:6 | glycerophospholipids | 25.30 | 8.91 | 5.98 |
| PC aa C42:0 | Phosphatidylcholine diacyl C42:0 | glycerophospholipids | 0.61 | 0.16 | 8.99 |
| PC aa C42:1 | Phosphatidylcholine diacyl C42:1 | glycerophospholipids | 0.32 | 0.08 | 11.89 |
| PC aa C42:2 | Phosphatidylcholine diacyl C42:2 | glycerophospholipids | 0.20 | 0.05 | 13.41 |
| PC aa C42:4 | Phosphatidylcholine diacyl C42:4 | glycerophospholipids | 0.18 | 0.05 | 14.39 |
| PC aa C42:5 | Phosphatidylcholine diacyl C42:5 | glycerophospholipids | 0.31 | 0.09 | 12.66 |
| PC aa C42:6 | Phosphatidylcholine diacyl C42:6 | glycerophospholipids | 0.40 | 0.11 | 12.18 |
| PC ae C30:0 | Phosphatidylcholine acyl-alkyl C30:0 | glycerophospholipids | 0.34 | 0.11 | 9.22 |
| PC ae C30:1 | Phosphatidylcholine acyl-alkyl C30:1 | glycerophospholipids | 0.19 | 0.09 | 39.59 |
| PC ae C30:2 | Phosphatidylcholine acyl-alkyl C30:2 | glycerophospholipids | 0.08 | 0.03 | 10.95 |
| PC ae C32:1 | Phosphatidylcholine acyl-alkyl C32:1 | glycerophospholipids | 8.52 | 2.15 | 7.08 |
| PC ae C32:2 | Phosphatidylcholine acyl-alkyl C32:2 | glycerophospholipids | 2.18 | 0.57 | 7.72 |
| PC ae C34:0 | Phosphatidylcholine acyl-alkyl C34:0 | glycerophospholipids | 3.48 | 1.08 | 9.04 |
| PC ae C34:1 | Phosphatidylcholine acyl-alkyl C34:1 | glycerophospholipids | 25.52 | 6.75 | 7.14 |
| PC ae C34:2 | Phosphatidylcholine acyl-alkyl C34:2 | glycerophospholipids | 33.53 | 9.75 | 6.37 |
| PC ae C34:3 | Phosphatidylcholine acyl-alkyl C34:3 | glycerophospholipids | 22.78 | 7.21 | 6.14 |
| PC ae C36:0 | Phosphatidylcholine acyl-alkyl C36:0 | glycerophospholipids | 1.46 | 0.43 | 12.87 |
| PC ae C36:1 | Phosphatidylcholine acyl-alkyl C36:1 | glycerophospholipids | 15.51 | 4.32 | 7.98 |
| PC ae C36:2 | Phosphatidylcholine acyl-alkyl C36:2 | glycerophospholipids | 29.32 | 8.47 | 6.47 |
| PC ae C36:3 | Phosphatidylcholine acyl-alkyl C36:3 | glycerophospholipids | 17.20 | 4.66 | 6.13 |
| PC ae C36:4 | Phosphatidylcholine acyl-alkyl C36:4 | glycerophospholipids | 38.84 | 11.28 | 6.06 |
| PC ae C36:5 | Phosphatidylcholine acyl-alkyl C36:5 | glycerophospholipids | 25.30 | 7.70 | 6.01 |
| PC ae C38:0 | Phosphatidylcholine acyl-alkyl C38:0 | glycerophospholipids | 2.92 | 0.93 | 7.10 |
| PC ae C38:1 | Phosphatidylcholine acyl-alkyl C38:1 | glycerophospholipids | 0.50 | 0.46 | 95.88 |
| PC ae C38:2 | Phosphatidylcholine acyl-alkyl C38:2 | glycerophospholipids | 2.78 | 0.88 | 15.76 |
| PC ae C38:3 | Phosphatidylcholine acyl-alkyl C38:3 | glycerophospholipids | 6.92 | 1.97 | 7.09 |
| PC ae C38:4 | Phosphatidylcholine acyl-alkyl C38:4 | glycerophospholipids | 22.59 | 5.63 | 6.00 |
| PC ae C38:5 | Phosphatidylcholine acyl-alkyl C38:5 | glycerophospholipids | 27.44 | 6.88 | 5.32 |
| PC ae C38:6 | Phosphatidylcholine acyl-alkyl C38:6 | glycerophospholipids | 11.04 | 3.11 | 5.72 |
| PC ae C40:1 | Phosphatidylcholine acyl-alkyl C40:1 | glycerophospholipids | 1.59 | 0.43 | 8.23 |
| PC ae C40:2 | Phosphatidylcholine acyl-alkyl C40:2 | glycerophospholipids | 1.99 | 0.53 | 7.29 |
| PC ae C40:3 | Phosphatidylcholine acyl-alkyl C40:3 | glycerophospholipids | 1.31 | 0.32 | 10.32 |
| PC ae C40:4 | Phosphatidylcholine acyl-alkyl C40:4 | glycerophospholipids | 2.91 | 0.70 | 6.29 |
| PC ae C40:5 | Phosphatidylcholine acyl-alkyl C40:5 | glycerophospholipids | 4.09 | 0.98 | 6.04 |
| PC ae C40:6 | Phosphatidylcholine acyl-alkyl C40:6 | glycerophospholipids | 5.42 | 1.50 | 6.15 |
| PC ae C42:0 | Phosphatidylcholine acyl-alkyl C42:0 | glycerophospholipids | 0.61 | 0.11 | 9.26 |
| PC ae C42:1 | Phosphatidylcholine acyl-alkyl C42:1 | glycerophospholipids | 0.43 | 0.10 | 10.62 |
| PC ae C42:2 | Phosphatidylcholine acyl-alkyl C42:2 | glycerophospholipids | 0.54 | 0.14 | 10.12 |
| PC ae C42:3 | Phosphatidylcholine acyl-alkyl C42:3 | glycerophospholipids | 0.78 | 0.20 | 8.37 |
| PC ae C42:4 | Phosphatidylcholine acyl-alkyl C42:4 | glycerophospholipids | 0.81 | 0.23 | 9.08 |
| PC ae C42:5 | Phosphatidylcholine acyl-alkyl C42:5 | glycerophospholipids | 2.02 | 0.50 | 6.50 |
| PC ae C44:3 | Phosphatidylcholine acyl-alkyl C44:3 | glycerophospholipids | 0.13 | 0.04 | 18.50 |
| PC ae C44:4 | Phosphatidylcholine acyl-alkyl C44:4 | glycerophospholipids | 0.30 | 0.09 | 15.16 |
| PC ae C44:5 | Phosphatidylcholine acyl-alkyl C44:5 | glycerophospholipids | 1.33 | 0.39 | 7.97 |
| PC ae C44:6 | Phosphatidylcholine acyl-alkyl C44:6 | glycerophospholipids | 1.01 | 0.29 | 7.95 |
| SM (OH) C14:1 | Hydroxysphingomyeline C14:1 | sphingolipids | 4.49 | 1.31 | 6.70 |
| SM (OH) C16:1 | Hydroxysphingomyeline C16:1 | sphingolipids | 1.89 | 0.54 | 6.82 |
| SM (OH) C22:1 | Hydroxysphingomyeline C22:1 | sphingolipids | 3.10 | 0.83 | 7.37 |
| SM (OH) C22:2 | Hydroxysphingomyeline C22:2 | sphingolipids | 2.85 | 0.79 | 7.42 |
| SM (OH) C24:1 | Hydroxysphingomyeline C24:1 | sphingolipids | 0.23 | 0.07 | 12.28 |
| SM C16:0 | Sphingomyeline C16:0 | sphingolipids | 63.10 | 13.77 | 5.84 |
| SM C16:1 | Sphingomyeline C16:1 | sphingolipids | 9.20 | 2.30 | 5.73 |
| SM C18:0 | Sphingomyeline C18:0 | sphingolipids | 10.07 | 2.59 | 6.33 |
| SM C18:1 | Sphingomyeline C18:1 | sphingolipids | 5.04 | 1.39 | 5.97 |
| SM C20:2 | Sphingomyeline C20:2 | sphingolipids | 0.15 | 0.06 | 20.48 |
| SM C24:0 | Sphingomyeline C24:0 | sphingolipids | 3.84 | 0.94 | 6.87 |
| SM C24:1 | Sphingomyeline C24:1 | sphingolipids | 8.73 | 2.14 | 7.71 |
| SM C26:0 | Sphingomyeline C26:0 | sphingolipids | 0.03 | 0.01 | 27.88 |
| SM C26:1 | Sphingomyeline C26:1 | sphingolipids | 0.05 | 0.02 | 26.15 |

#### Table S6. Overview of the 148 plasma proteins quantified using Scanning SWATH in the CHRIS cohort.

| **Protein** | **Gene** | **Description** | **Mean** | **SD** | **CV QC (%)** |
| --- | --- | --- | --- | --- | --- |
| P02768 | *ALB* | Serum albumin | 16.64 | 0.20 | 9.47 |
| P02766 | *TTR* | Transthyretin | 11.26 | 0.57 | 20.03 |
| P19827 | *ITIH1* | Inter-alpha-trypsin inhibitor heavy chain H1 | 13.36 | 0.21 | 6.75 |
| P01023 | *A2M* | Alpha-2-macroglobulin | 13.72 | 0.42 | 7.06 |
| P01042;P01042-2 | *KNG1* | Kininogen-1 | 13.06 | 0.21 | 8.42 |
| P02649 | *APOE* | Apolipoprotein E | 12.75 | 0.48 | 13.78 |
| P01024 | *C3* | Complement C3 | 13.88 | 0.20 | 4.87 |
| P04196 | *HRG* | Histidine-rich glycoprotein | 12.63 | 0.39 | 9.18 |
| P01011 | *SERPINA3* | Alpha-1-antichymotrypsin | 13.49 | 0.28 | 10.99 |
| P02787 | *TF* | Serotransferrin | 14.26 | 0.25 | 7.59 |
| P01834 | *IGKC* | Immunoglobulin kappa constant | 13.74 | 0.32 | 7.62 |
| Q14624;Q14624-2 | *ITIH4* | Inter-alpha-trypsin inhibitor heavy chain H4 | 12.98 | 0.22 | 7.54 |
| P07360 | *C8G* | Complement component C8 gamma chain | 11.61 | 0.43 | 18.81 |
| P00450 | *CP* | Ceruloplasmin | 12.85 | 0.35 | 8.33 |
| P02647 | *APOA1* | Apolipoprotein A-I | 16.26 | 0.41 | 40.41 |
| P02760 | *AMBP* | Protein AMBP | 14.50 | 0.22 | 8.80 |
| P01008 | *SERPINC1* | Antithrombin-III | 13.24 | 0.20 | 7.37 |
| O95445 | *APOM* | Apolipoprotein M | 14.07 | 0.35 | 14.76 |
| P01031 | *C5* | Complement C5 | 11.91 | 0.22 | 8.02 |
| P01042-2 | *KNG1* | Isoform LMW of Kininogen-1 | 13.30 | 0.41 | 17.43 |
| P06396;P06396-2 | *GSN* | Gelsolin | 12.16 | 0.29 | 10.48 |
| P25311 | *AZGP1* | Zinc-alpha-2-glycoprotein | 12.58 | 0.27 | 9.66 |
| P22792 | *CPN2* | Carboxypeptidase N subunit 2 | 11.28 | 0.49 | 21.87 |
| Q96PD5 | *PGLYRP2* | N-acetylmuramoyl-L-alanine amidase | 12.30 | 0.38 | 11.70 |
| P19823 | *ITIH2* | Inter-alpha-trypsin inhibitor heavy chain H2 | 14.08 | 0.20 | 8.58 |
| P02675 | *FGB* | Fibrinogen beta chain | 14.11 | 0.80 | 29.20 |
| P02748 | *C9* | Complement component C9 | 12.01 | 0.39 | 9.33 |
| P05155;P05155-3 | *SERPING1* | Plasma protease C1 inhibitor | 12.68 | 0.93 | 16.47 |
| P43652 | *AFM* | Afamin | 12.82 | 0.31 | 9.57 |
| P02679;P02679-2 | *FGG* | Fibrinogen gamma chain | 14.88 | 0.75 | 27.52 |
| P01861 | *IGHG4* | Immunoglobulin heavy constant gamma 4 | 12.57 | 1.16 | 12.48 |
| P06727 | *APOA4* | Apolipoprotein A-IV | 12.90 | 0.43 | 7.04 |
| O14791;O14791-2;O14791-3 | *APOL1* | Apolipoprotein L1 | 11.68 | 0.46 | 14.86 |
| P00751 | *CFB* | Complement factor B | 12.41 | 0.24 | 7.50 |
| P02750 | *LRG1* | Leucine-rich alpha-2-glycoprotein | 12.71 | 0.43 | 10.84 |
| P20851;P20851-2 | *C4BPB* | C4b-binding protein beta chain | 13.27 | 0.48 | 22.64 |
| P13671 | *C6* | Complement component C6 | 12.71 | 0.32 | 14.02 |
| P01019 | *AGT* | Angiotensinogen | 13.45 | 0.66 | 10.60 |
| P35858;P35858-2 | *IGFALS* | Insulin-like growth factor-binding protein complex acid labile subunit | 11.87 | 0.45 | 20.12 |
| P02671 | *FGA* | Fibrinogen alpha chain | 14.43 | 0.83 | 31.07 |
| P04114 | *APOB* | Apolipoprotein B-100 | 12.52 | 0.41 | 9.90 |
| P36955 | *SERPINF1* | Pigment epithelium-derived factor | 11.52 | 0.30 | 13.08 |
| P07357 | *C8A* | Complement component C8 alpha chain | 12.52 | 0.28 | 12.18 |
| P02751 | *FN1* | Fibronectin | 11.60 | 0.94 | 61.10 |
| P02765 | *AHSG* | Alpha-2-HS-glycoprotein | 12.77 | 0.28 | 9.31 |
| P00488 | *F13A1* | Coagulation factor XIII A chain | 10.90 | 0.67 | 30.23 |
| A0A0C4DH29 | *IGHV1-3* | Immunoglobulin heavy variable 1-3 | 10.67 | 0.67 | 42.75 |
| P01602 | *IGKV1-5* | Immunoglobulin kappa variable 1-5 | 12.53 | 0.54 | 14.57 |
| P01619 | *IGKV3-20* | Immunoglobulin kappa variable 3-20 | 12.80 | 0.53 | 16.85 |
| P10909;P10909-2;P10909-4;P10909-5 | *CLU* | Clusterin | 12.86 | 0.16 | 6.27 |
| P00746 | *CFD* | Complement factor D | 10.40 | 0.56 | 26.58 |
| P15814 | *IGLL1* | Immunoglobulin lambda-like polypeptide 1 | 13.73 | 0.46 | 23.85 |
| P00747 | *PLG* | Plasminogen | 13.36 | 0.22 | 6.52 |
| P08571 | *CD14* | Monocyte differentiation antigen CD14 | 9.58 | 0.60 | 36.80 |
| P02749 | *APOH* | Beta-2-glycoprotein 1 | 12.95 | 0.42 | 18.55 |
| P04217;P04217-2 | *A1BG* | Alpha-1B-glycoprotein | 13.22 | 0.26 | 10.61 |
| P00738 | *HP* | Haptoglobin | 14.76 | 0.63 | 10.07 |
| P06681 | *C2* | Complement C2 | 11.59 | 0.40 | 21.37 |
| P05543 | *SERPINA7* | Thyroxine-binding globulin | 10.48 | 0.52 | 18.28 |
| P01009 | *SERPINA1* | Alpha-1-antitrypsin | 14.28 | 0.31 | 7.95 |
| P04004 | *VTN* | Vitronectin | 13.34 | 0.26 | 8.55 |
| P05154 | *SERPINA5* | Plasma serine protease inhibitor | 10.18 | 0.80 | 39.75 |
| P01859 | *IGHG2* | Immunoglobulin heavy constant gamma 2 | 16.03 | 0.47 | 8.07 |
| P08603 | *CFH* | Complement factor H | 11.99 | 0.23 | 7.93 |
| P04003 | *C4BPA* | C4b-binding protein alpha chain | 12.59 | 0.30 | 8.64 |
| P01860 | *IGHG3* | Immunoglobulin heavy constant gamma 3 | 13.75 | 0.66 | 17.12 |
| P02790 | *HPX* | Hemopexin | 13.85 | 0.20 | 7.89 |
| P01591 | *JCHAIN* | Immunoglobulin J chain | 11.23 | 0.62 | 16.03 |
| P02656 | *3* | Apolipoprotein C-III | 14.56 | 0.43 | 9.07 |
| P01877 | *IGHA2* | Immunoglobulin heavy constant alpha 2 | 14.42 | 0.65 | 11.79 |
| P01876 | *IGHA1* | Immunoglobulin heavy constant alpha 1 | 15.90 | 0.67 | 13.14 |
| P01857 | *IGHG1* | Immunoglobulin heavy constant gamma 1 | 15.83 | 0.37 | 12.26 |
| P01871 | *IGHM* | Immunoglobulin heavy constant mu | 14.59 | 0.76 | 10.95 |
| P00734 | *F2* | Prothrombin | 12.28 | 0.20 | 7.02 |
| A0A0J9YX35 | *IGHV3-64D* | Immunoglobulin heavy variable 3-64D | 12.44 | 0.53 | 19.31 |
| P02753 | *RBP4* | Retinol-binding protein 4 | 13.20 | 0.34 | 9.48 |
| P02746 | *C1QB* | Complement C1q subcomponent subunit B | 12.54 | 0.33 | 13.13 |
| P08697 | *SERPINF2* | Alpha-2-antiplasmin | 12.40 | 0.27 | 12.31 |
| P06310 | *IGKV2-30* | Immunoglobulin kappa variable 2-30 | 12.42 | 0.58 | 16.24 |
| P0C0L4 | *C4A* | Complement C4-A | 12.60 | 0.70 | 18.79 |
| P03952 | *KLKB1* | Plasma kallikrein | 11.17 | 0.32 | 13.15 |
| P07358 | *C8B* | Complement component C8 beta chain | 12.02 | 0.36 | 17.69 |
| P02654 | *APOC1* | Apolipoprotein C-I | 13.65 | 0.79 | 31.39 |
| P02774;P02774-3 | *GC* | Vitamin D-binding protein | 13.95 | 0.22 | 6.13 |
| P68871 | *HBB* | Hemoglobin subunit beta | 14.19 | 0.48 | 17.13 |
| P23142 | *FBLN1* | Fibulin-1 | 10.83 | 0.69 | 34.48 |
| Q9HCU4 | *CELSR2* | Cadherin EGF LAG seven-pass G-type receptor 2 | 14.07 | 1.12 | 45.74 |
| Q16610;Q16610-4 | *ECM1* | Extracellular matrix protein 1 | 11.61 | 0.50 | 23.91 |
| P02652 | *APOA2* | Apolipoprotein A-II | 16.55 | 0.22 | 8.17 |
| P00748 | *F12* | Coagulation factor XII | 13.45 | 0.47 | 11.65 |
| P27169 | *PON1* | Serum paraoxonase/arylesterase 1 | 12.14 | 0.44 | 11.32 |
| P80108 | *GPLD1* | Phosphatidylinositol-glycan-specific phospholipase D | 10.42 | 0.43 | 18.47 |
| P51884 | *LUM* | Lumican | 11.31 | 0.39 | 14.57 |
| P02747 | *C1QC* | Complement C1q subcomponent subunit C | 13.75 | 0.32 | 10.57 |
| A0A075B6I0 | *IGLV8-61* | Immunoglobulin lambda variable 8-61 | 12.74 | 0.79 | 15.60 |
| A0A075B6H9 | *IGLV4-69* | Immunoglobulin lambda variable 4-69 | 10.90 | 0.79 | 34.15 |
| P05546 | *SERPIND1* | Heparin cofactor 2 | 12.87 | 0.35 | 7.99 |
| P22352 | *GPX3* | Glutathione peroxidase 3 | 11.08 | 0.49 | 23.44 |
| P06396 | *GSN* | Gelsolin | 13.10 | 0.53 | 28.30 |
| P09871 | *C1S* | Complement C1s subcomponent | 11.83 | 0.27 | 11.64 |
| P05156 | *CFI* | Complement factor I | 12.40 | 0.29 | 11.31 |
| A0A0B4J1U7 | *IGHV6-1* | Immunoglobulin heavy variable 6-1 | 15.42 | 1.28 | 15.19 |
| A0A0J9YXX1 | *IGHV5-10-1* | Immunoglobulin heavy variable 5-10-1 | 13.54 | 1.44 | 19.24 |
| P01780 | *IGHV3-7* | Immunoglobulin heavy variable 3-7 | 13.03 | 0.64 | 23.46 |
| A0A0A0MS15 | *IGHV3-49* | Immunoglobulin heavy variable 3-49 | 13.68 | 0.56 | 23.31 |
| P00736 | *C1R* | Complement C1r subcomponent | 12.16 | 0.30 | 15.11 |
| Q06033;Q06033-2 | *ITIH3* | Inter-alpha-trypsin inhibitor heavy chain H3 | 11.89 | 0.57 | 24.09 |
| P08519 | *LPA* | Apolipoprotein(a) | 11.75 | 1.87 | 56.51 |
| P08185 | *SERPINA6* | Corticosteroid-binding globulin | 13.53 | 0.57 | 17.06 |
| P04217 | *A1BG* | Alpha-1B-glycoprotein | 13.33 | 0.23 | 9.36 |
| P04278 | *SHBG* | Sex hormone-binding globulin | 11.10 | 1.25 | 34.72 |
| P15169 | *CPN1* | Carboxypeptidase N catalytic chain | 10.91 | 0.43 | 20.21 |
| P05090 | *APOD* | Apolipoprotein D | 12.00 | 0.40 | 13.12 |
| P10643 | *C7* | Complement component C7 | 11.73 | 0.41 | 17.89 |
| Q14624 | *ITIH4* | Inter-alpha-trypsin inhibitor heavy chain H4 | 11.92 | 0.32 | 15.29 |
| P02763 | *ORM1* | Alpha-1-acid glycoprotein 1 | 14.97 | 0.43 | 11.61 |
| P20742 | *PZP* | Pregnancy zone protein | 13.21 | 0.84 | 31.92 |
| P18428 | *LBP* | Lipopolysaccharide-binding protein | 10.81 | 0.76 | 37.90 |
| O43866 | *CD5L* | CD5 antigen-like | 12.22 | 0.75 | 17.79 |
| P02745 | *C1QA* | Complement C1q subcomponent subunit A | 12.75 | 0.46 | 24.05 |
| P07225 | *PROS1* | Vitamin K-dependent protein S | 11.23 | 0.31 | 14.38 |
| P29622 | *SERPINA4* | Kallistatin | 11.69 | 0.49 | 16.81 |
| O75636 | *FCN3* | Ficolin-3 | 12.17 | 0.60 | 17.67 |
| P01701 | *IGLV1-51* | Immunoglobulin lambda variable 1-51 | 12.48 | 0.78 | 30.98 |
| P01703 | *IGLV1-40* | Immunoglobulin lambda variable 1-40 | 11.47 | 0.88 | 35.82 |
| P06312 | *IGKV4-1* | Immunoglobulin kappa variable 4-1 | 11.34 | 0.58 | 22.12 |
| P01705 | *IGLV2-23* | Immunoglobulin lambda variable 2-23 | 11.73 | 0.84 | 29.62 |
| P49908 | *SELENOP* | Selenoprotein P | 11.07 | 0.59 | 29.36 |
| O75882-2 | *ATRN* | Isoform 2 of Attractin | 11.87 | 0.34 | 17.01 |
| Q9UGM5 | *FETUB* | Fetuin-B | 11.81 | 0.87 | 30.66 |
| P05452 | *CLEC3B* | Tetranectin | 12.78 | 0.32 | 13.91 |
| A0A0B4J1V2 | *IGHV2-26* | Immunoglobulin heavy variable 2-26 | 13.31 | 0.68 | 15.91 |
| P00742 | *F10* | Coagulation factor X | 12.81 | 0.32 | 16.85 |
| P00739 | *HPR* | Haptoglobin-related protein | 11.94 | 0.61 | 11.90 |
| P01599 | *IGKV1-17* | Immunoglobulin kappa variable 1-17 | 12.56 | 0.78 | 22.94 |
| P00740 | *F9* | Coagulation factor IX | 11.19 | 0.81 | 30.10 |
| P19652 | *ORM2* | Alpha-1-acid glycoprotein 2 | 14.04 | 0.44 | 10.02 |
| P05160 | *F13B* | Coagulation factor XIII B chain | 10.87 | 0.66 | 30.77 |
| A0A075B6K4 | *IGLV3-10* | Immunoglobulin lambda variable 3-10 | 11.19 | 0.94 | 38.84 |
| A0A0C4DH31 | *IGHV1-18* | Immunoglobulin heavy variable 1-18 | 11.63 | 0.81 | 36.85 |
| P80748 | *IGLV3-21* | Immunoglobulin lambda variable 3-21 | 12.51 | 0.79 | 34.45 |
| A0A0B4J1Y9 | *IGHV3-72* | Immunoglobulin heavy variable 3-72 | 11.02 | 0.85 | 50.65 |
| P23142;P23142-4 | *FBLN1* | Fibulin-1 | 12.16 | 0.59 | 22.00 |
| P69905 | *HBA1* | Hemoglobin subunit alpha | 14.20 | 0.52 | 20.06 |
| A0A075B6J9 | *IGLV2-18* | Immunoglobulin lambda variable 2-18 | 10.54 | 0.99 | 34.59 |
| A0A0C4DH34 | *IGHV4-28* | Immunoglobulin heavy variable 4-28 | 11.63 | 0.66 | 22.39 |
| B9A064 | *IGLL5* | Immunoglobulin lambda-like polypeptide 5 | 16.33 | 0.43 | 13.59 |
| P06276 | *BCHE* | Cholinesterase | 10.84 | 0.62 | 34.76 |

### Supplementary Figures

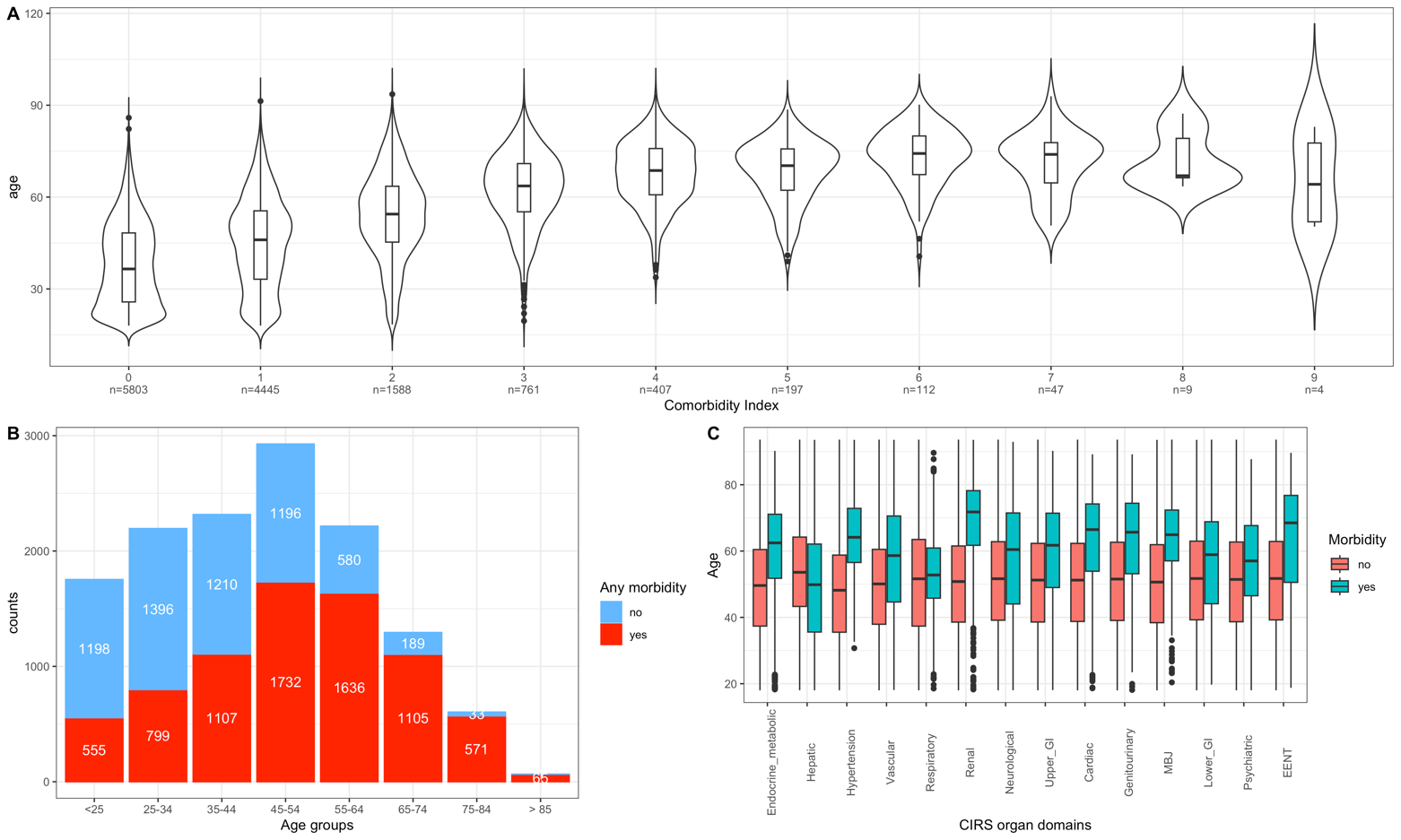

Figure S1. Distribution of age and health status in the CHRIS cohort. **A)** Age distribution by the Comorbidity Index, which is expressed as the total number of CIRS organ domains scoring ≥2. **B)** Absolute distribution of “Any morbidity” (yes+no) by age-group. The relative proportion of red-bars within each stacked column-bar represents the age-group specific prevalence of any morbidity. **C)** Age distribution by morbidity conditions in the specific CIRS domains.

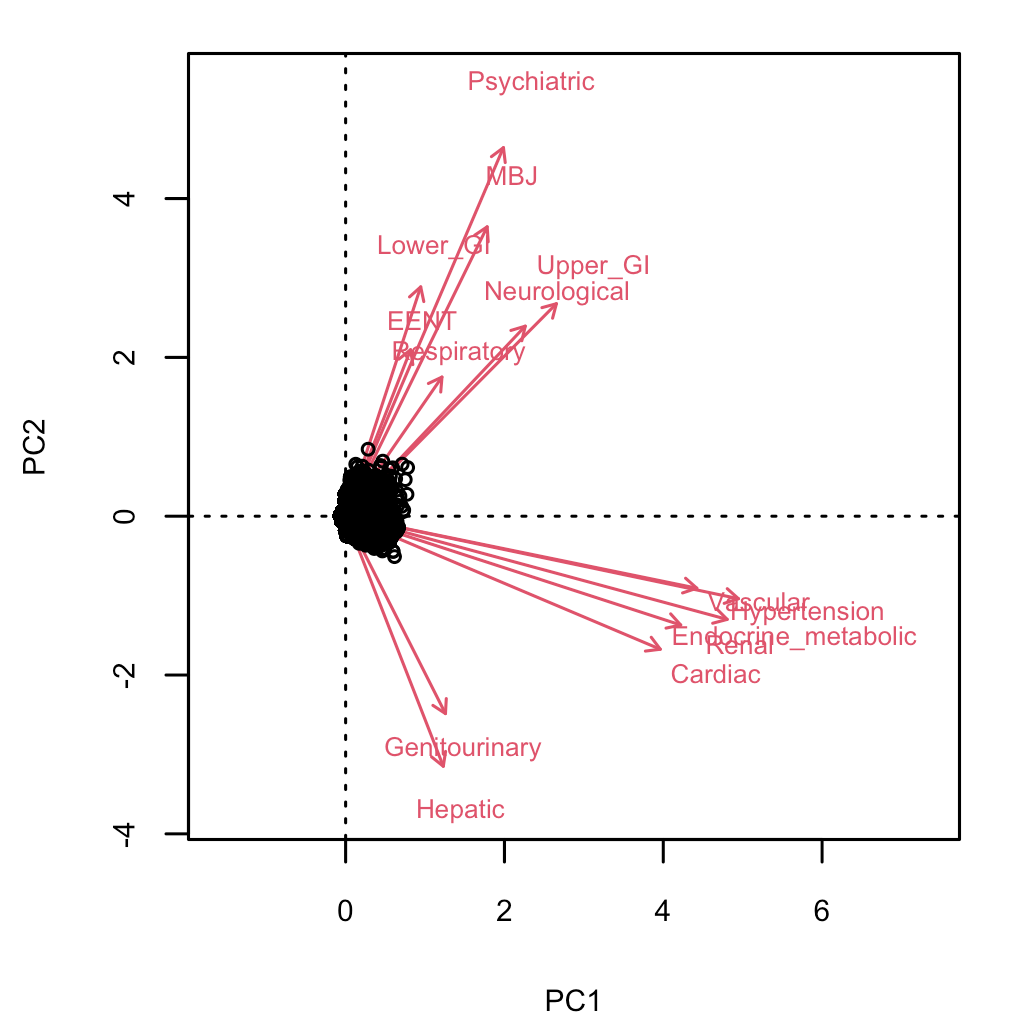

Figure S2. Biplot of ordinary correspondence analysis of comorbidities in the CHRIS cohort. CIRS domains with a stronger relation have longer (size consistency) and closer (direction consistency) loadings.

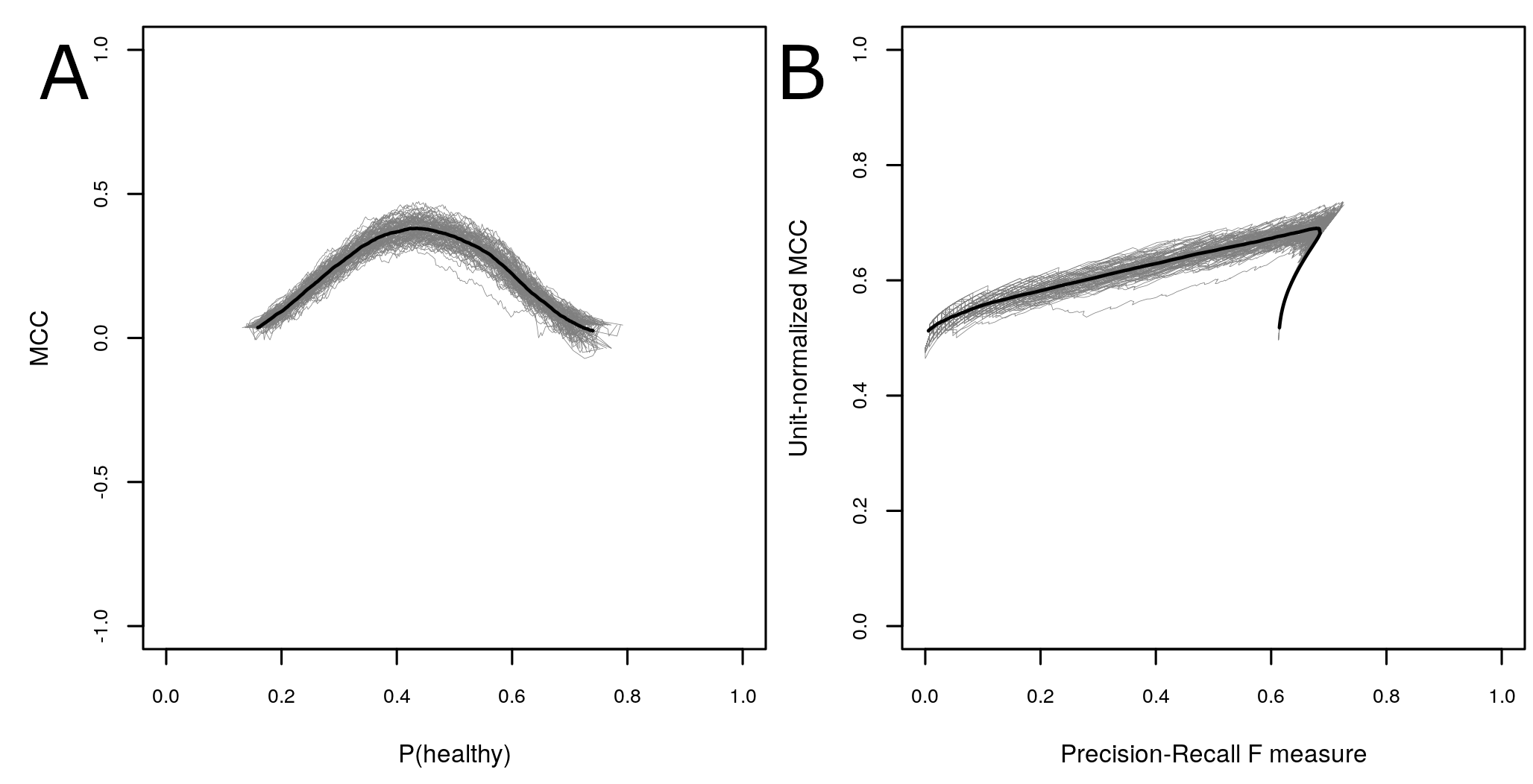

Figure S3. Performance evaluation of 100 random forest models including as predictors age, sex, 174 metabolites and 148 proteins to classify health status (any morbidity vs. healthy). The performance is expressed by A) MCC and B) MCC-F1. In the plots each validation run is represented as a gray line and the averaged curves are shown as a thicker black line.

### Supplementary Text

#### Text S1. Description of Random Forest Performance Evaluation Scores and model comparison.

**Random Forest Performance Evaluation Scores**

We make use of the following measures:

- AUC: area under the receiver operating characteristic (ROC) curve.
- Precision: positive predictive value;
- F1: F1 score, which is the harmonic mean between precision and sensitivity. i.e. F1 = (precision x TPR) / (precision + TPR);
- MCC. the Matthews correlation coefficient. which uses absolute numbers instead of ratios. MCC = 0 corresponds to a pure chance guess. TP, true positive; TN, true negative; FP, false positive; FN, false negative.

$$MCC =\frac{TP \times TN - FP \times FN}{\sqrt{(TP + FP) \times(TP + FN) \times(TN + FP) \times(TN + FN)}}$$

- MCCF1. the unit-normalized Matthews correlation coefficient (MCC+1)/2. MCCF1 = 0.5 corresponds to the chance event.

**Random forest model comparison.**

For the RF analysis we built different models that included varying sets of the predictors, which were a) age and sex, b) age, sex and metabolites, c) age, sex and proteins, and d) age, sex, metabolites, proteins, and compared the performance across all models.

In general, model performance increased by adding metabolites and proteins to age and sex, whereas addition of proteins alone did not improve model performance. An overview of the performance of all models is provided in **Table S7** below.

| **Table S7. Overview of performance summary measures for the random forest models.** The calculated mean from the 100 model performance evaluations on the respective test set and the 95% confidence intervals (95% CI). | | | | |
| --- | --- | --- | --- | --- |
| **Model** | **TPR** | **FPR** | **AUC** | **MCC** |
| a) age + sex | 0.706 (0.697.0.715) | 0.320 (0.312.0.329) | 0.723 (0.719.0.727) | 0.385(0.378.0.391)) |
| b) age + sex + mbx | 0.750 (0.737.0.763) | 0.351 (0.338.0.364) | 0.748 (0.744.0.752) | 0.400 (0.394.0.406) |
| c) age + sex + ptx | 0.749 (0.739.0.759) | 0.338 (0.329.0.348) | 0.740 (0.736.0.744) | 0.410 (0.404.0.416) |
| d) age + sex + mbx + ptx | 0.740 (0.729.0.750) | 0.342 (0.332.0.352) | 0.747 (0.743.0.751) | 0.397 (0.391.0.404) |
| TPR = true positive rate (or sensitivity). FPR = false positive rate (or specificity). AUC = area under the receiver operating characteristic (ROC) curve. MCC = Matthew’s correlation coefficient. MCC = 0 corresponds to a pure chance guess | | | | |

To evaluate whether there was a statistically significant (p-value<0.05) difference in the performance, we compared the MCC values standardized to estimate effect sizes and implemented the Student’s t-test. A statistically significant (*p*-value<0.05) improvement was only observed from models b), c) and d) to model a) but not between models b), c) and d) (**see Figure S6 below**). Given this observation and the similarity of the performance for models d) and b), we decided to focus the proceeding analyses on the full model d).

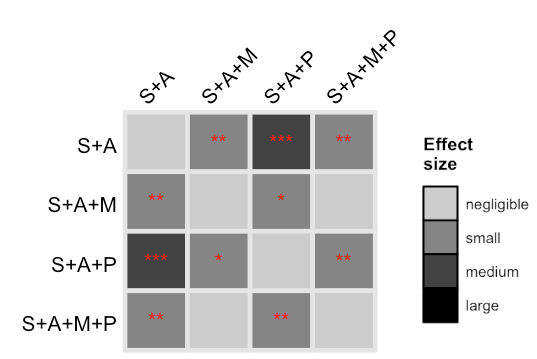

**Figure S4. Assessment of RF model performance comparing the standardized MCCs across the different predictor sets.** In the following plots we have categorized the effect sizes by magnitude: 0≤ s <0.2. negligible; 0.2≤ s<0.5. small; 0.5≤ s < 0.8; medium; and s ≥ 0.8. large. Comparisons of the effect sizes (standardized MCCs) for different predictor sets are grouped by health status (any morbidity vs. healthy). Predictor sets are the following: S+A. sex and age; S+A+M. sex. age. metabolites; S+A+P. sex. age. proteins; S+A+M+P. sex. age. metabolites. proteins. For each comparison. we performed a two-sided Student’s t-test. and asterisks on top of the effect size cells indicate the level of significance: no asterisk. P≥0.05; *. 0.01≤P<0.05; **. 0.001≤P<0.01; ***. P<0.001.

An overview of all significant results for the RF analyses for the different predictor sets is presented in **Figure** **S5** below**.**

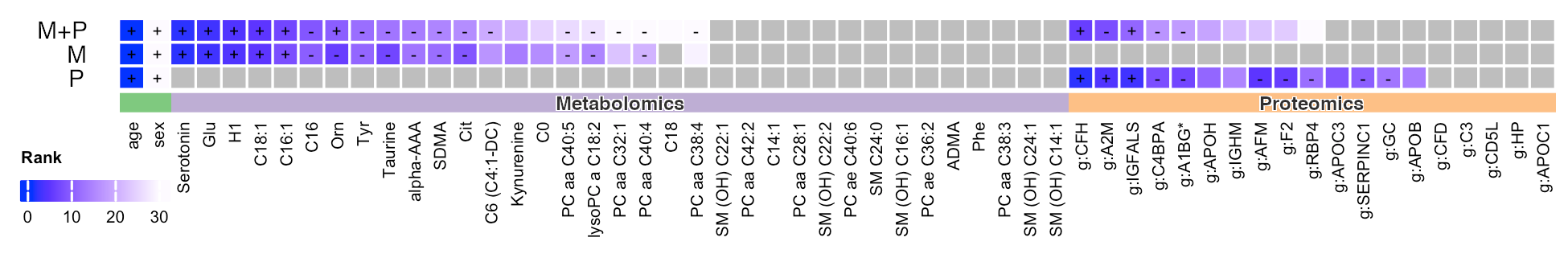

**Figure S5. Overview of drop in Gini index for most important predictor sets.** The models included only metabolites (M). only proteins (P) and the combined predictors (M+P). healthy:amorb. any morbidity vs healthy. Predictors are marked by the number of times a *p*-value was significant in the permutation runs for such a variable (*p*<0.05): +. all 100 runs resulted a significant *p*-value; -. between 80 and 99 runs were associated with significant *p*-values; no sign. between 50 and 79 runs were associated with significant *p*-values. Results are restricted to variables that are significant in at least 50% of all permutation runs.

#### Text S2. Discussion of additional features

In this section we are discussing relevant molecular features that were determined with the RF model, where the age-independent associations were confirmed by linear regression, but for which the difference in abundances was lower than the coefficient of variation cut-off

Overall, twelve such features were identified (hexose, C18:1, C16:1, lyso PC a C18:2, PC aa C32:1, tyrosine, taurine, kynurenine, AFM, CFH, RBP4, A1BG). Hexoses have been previously associated with metabolic health ^1,2^. Several long-chain glycerophospholipids and long-chain acylcarnitines were also related to health status. Acylcarnitine levels are known to increase with age, as they accumulate because of incompletely oxidized fatty acids in the mitochondria beta oxidation pathway ^3,4^. More specifically, circulating long-chain acylcarnitine concentrations are indicators of mitochondrial functionality, with increased levels indicating reduced oxidative phosphorylation capacity ^5^ and previous studies reported an increase of carnitines in cardiometabolic disease and cancer ^5–7^. Tyrosine, kynurenine and taurine have been associated with increased risk of type 2 diabetes ^8^, mortality ^9^, metabolic syndrome, neurodegeneration and frailty ^10–13^. Taurine is involved in mitochondrial function, apoptosis and serves as an anti-oxidant showing anti-inflammatory properties ^14^. Afamin (AFM) has been reported to play a role in vitamin E transport and higher afamin levels are associated with poor metabolic health ^15–18^. Tanaka et al further reported associations between AFM and mortality ^19^. Retinol-binding protein 4 (RBP4) is a specific carrier for retinol in the blood and has been implicated in metabolic health and inflammation ^20–22^. The complement factor H (CFH) plays a key role in the regulation of alternative pathway of complement system ^23^. Alpha-1B-glycoprotein (A1BG) has been reported as potential urinary biomarker for chronic kidney disease ^24^.
